# A MULTI-MINERAL INTERVENTION TO MODULATE COLONIC MUCOSAL PROTEIN PROFILE: Results from a 90-day trial in healthy human subjects

**DOI:** 10.1101/2020.12.30.20249070

**Authors:** Muhammad N. Aslam, Shannon D. McClintock, Mohamed Ali H. Jawad-Makki, Karsten Knuver, Haris M. Ahmad, Venkatesha Basrur, Ingrid L. Bergin, Suzanna M. Zick, Ananda Sen, D. Kim Turgeon, James Varani

**Affiliations:** Departments of Pathology, The University of Michigan School of Public Health, Ann Arbor, MI 48109; The Unit for Laboratory Animal Medicine, The University of Michigan Medical School, Ann Arbor, MI 48109; Departments of Family Medicine, The University of Michigan School of Public Health, Ann Arbor, MI 48109; Department of Nutritional Science, The University of Michigan School of Public Health, Ann Arbor, MI 48109; Departments of Biostatistics, The University of Michigan School of Public Health, Ann Arbor, MI 48109; Departments of Internal Medicine (Division of Gastroenterology), The University of Michigan School of Public Health, Ann Arbor, MI 48109

**Keywords:** Aquamin^®^, biomarkers, calcium, colon cancer chemoprevention, proteomic analysis

## Abstract

The overall goal of this study was to determine if Aquamin^®^, a calcium- and magnesium-rich natural product, would alter the expression of proteins involved in growth-regulation, differentiation and barrier formation in the colon. Thirty healthy human subjects were enrolled in a three-arm, 90-day interventional trial in which Aquamin^®^ (provided daily to deliver 800-mg of calcium per day) was compared to calcium alone and placebo. Before and after the 90-day interventional period, colonic biopsies were obtained. Biopsies were evaluated by immunohistology for expression of Ki67 (a proliferation marker) and for CK20 and p21 (differentiation markers). Tandem mass tag-mass spectrometry-based detection was used to assess levels of multiple proteins. As compared to placebo or calcium, Aquamin^®^ reduced the level of Ki67 expression (20%). Neither intervention altered CK20 expression, while a trend toward increased p21 was observed with calcium and Aquamin^®^ (117% and 99% respectively). In the proteomic screen, Aquamin^®^ treatment resulted in many more proteins being upregulated or downregulated (1.5 fold-change with ≤2% false-discovery rate) than placebo. Included among the upregulated proteins were cytokeratins, cell-cell adhesion molecules and components of the basement membrane. Many of the downregulated proteins were those involved in proliferation and nucleic acid metabolism. Calcium alone also altered the expression of many of the same proteins but not to the same extent as Aquamin^®^. We conclude that daily Aquamin^®^ ingestion alters protein expression profile in the colon that could be beneficial to colonic health. These data warrant additional studies with a larger sample size to validate these findings.

**Prevention Relevance:** A multi-mineral approach reduced proliferation and induced differentiation in *ex vivo* settings and has been shown to decrease colon polyp incidence in mouse (polyp-prevention) studies. The findings from a 90-day trial in human subjects (presented here) demonstrated improved biomarker-modulation efficacy, warranting to conduct the polyp-prevention trial in at-risk human subjects.

## INTRODUCTION

It is well-accepted that an adequate intake of dietary calcium is important for the prevention of several long-latency diseases [1-5] including colon cancer [6-11]. While calcium (as a single mineral) has been widely studied, it is now becoming recognized that other minerals along with calcium can provide benefit not obtainable with calcium alone [12-14]. Some of these (e.g., chromium, copper, iron, magnesium, manganese, molybdenum, potassium, selenium, sodium and zinc) have recommended dietary intake levels [15-19], but others are utilized in such small amounts that intake levels have not been defined. Past studies in our laboratory have made use of a multi-mineral natural product (Aquamin^®^) as part of a strategy for assessing the role of minerals in the prevention of long-latency diseases. Aquamin^®^ is derived from the mineralized remains of red marine algae and consists of calcium and magnesium along with detectable levels of 72 additional trace elements [20-27]. In regard to colon cancer, specifically, we have shown that Aquamin^®^ reduces proliferation and increases differentiation of human colon carcinoma cells in monolayer culture [20, 21] and in organoid culture [22-25] more effectively than calcium alone. Along with these *in vitro* studies, two long-term (15-18 month) studies in mice demonstrated the improved efficacy of Aquamin^®^ relative to that of calcium alone for inhibition of colon polyp formation [26, 27].

While these findings provide evidence for efficacy of the multi-mineral approach to disease prevention, they do not indicate whether a multi-mineral supplement such as Aquamin^®^ can be used in humans or whether the same beneficial activities observed in preclinical models will be seen in humans. To begin addressing this issue, we carried out a 90-day, FDA-approved pilot study, comparing daily intervention with Aquamin^®^ to calcium alone and placebo in healthy human subjects at increased risk for colorectal cancer (ClinicalTrials.gov Identifier NCT02647671). While the primary purpose of the pilot study was to assess safety and tolerability, we also evaluated microbial community structure and assessed bile acid, short-chain fatty acid (SFCA) and eicosanoid profiles at endpoint in comparison to baseline values in subjects from all three groups. Those data have been published recently [28]. As part of the same study, replicate colonic biopsies were obtained from each subject prior to intervention and at the end of the 90-day treatment period. One set of biopsies was evaluated morphometrically for histological changes (crypt length) and by quantitative immunohistology for expression of Ki67 as a proliferation marker, and for expression of CK20 and p21 as indicators of differentiation. These biomarkers of risk for colorectal cancer (CRC) have been used in past colon polyp prevention studies [19, 29-31]. Additional biopsies from each cohort of individuals were subjected to proteomic profile evaluation via tandem mass tag (TMT) labeled mass spectrometry. The results of the histological / immunohistological and proteomic studies are described in the current report. Findings from the interventional trial are compared to recent findings obtained using similar approaches in colon organoid culture [22-24].

## MATERIALS AND METHODS

### Interventions

Three study interventions were used, one experimental (Aquamin^®^) and two comparators (Calcium carbonate and maltodextrin). Aquamin^®^ is a natural product obtained from the skeletal remains of red marine algae of the *Lithothamnion* genus. Aquamin^®^ is approximately 30% calcium (by weight), contains calcium and magnesium in an approximately 12:1 ratio and has detectable levels of 72 additional trace minerals (essentially all of the minerals accumulated by the algae from seawater). Aquamin^®^ is sold as a food supplement (GRAS 000028) (Marigot Ltd, Cork, Ireland) and is used in various products for human consumption in Europe, Asia, Australia, and North America. A single batch of Aquamin-Food Grade^®^ was used for this study. The mineral composition was established via an independent laboratory (Advanced Laboratories; Salt Lake City, Utah) using Inductively Coupled Plasma - Optical Emission Spectrometry (*ICP***-***OES*). A complete list of elements detected in this batch of Aquamin^®^ and their relative amounts can be found in our recent report [28]. Calcium carbonate was used as an active comparator and maltodextrin was used as placebo. Daily intake of Aquamin^®^ or calcium alone was designed to provide 800 mg of calcium per day.

### Trial design

The design of this pilot study was described in detail in our earlier publication [28]. Briefly, this was a placebo-controlled, double-blind, trial in which thirty subjects were included. Thirty subjects were randomized into three arms, allocating 10 subjects per arm. These subjects were 18 to 80 years aged, male or non-pregnant female. Subjects were in general good health but with “an increased risk for colon cancer” based on a personal history of i) colorectal polyp, ii) early stage (stage I or II) colon cancer surgically removed without administration of adjuvant therapy, iii) stage III colon cancer treated with surgery more than 5 years ago or having a first-degree blood relative diagnosed with colorectal cancer under the age of 60 years. Exclusion criteria included history of kidney disease or kidney stones, Crohn’s disease or ulcerative colitis, gastrointestinal hemorrhagic disorders, or coagulopathy, hereditary non-polyposis coli or familial adenomatous polyposis.

This FDA-approved interventional trial was conducted at Michigan Medicine under an Investigational New Drug (IND) #118194. The Institutional Review Board at the University of Michigan Medical School (IRBMED) provided the required oversight. The study was listed on Clinicaltrials.gov (study identifier NCT02647671). All subjects provided written informed consent before participation. This phase I interventional trial involving human subjects was conducted in accordance with recognized ethical guidelines, for example, Declaration of Helsinki, International Ethical Guidelines for Biomedical Research Involving Human Subjects (CIOMS), the Belmont Report and the U.S. Common Rule.

Subjects underwent flexible sigmoidoscopy procedure at baseline. No bowel cleansing was performed prior to the procedure. Twelve 2.5 mm colonic mucosal biopsies were obtained using endoscopic pinch biopsy along with two stool specimens from within the sigmoid colon (20 cm above the anus). After baseline sigmoidoscopy, subjects were randomized to one of three groups. Ten subjects were given Aquamin capsules for 90 days, providing 800 mg of calcium per day. Ten subjects received 800 mg of calcium carbonate daily and ten subjects received maltodextrin as placebo. At the end of the 90-day intervention period (90±5 days), subjects again underwent unpreped flexible sigmoidoscopy. Eight colonic mucosal biopsies along with two stool specimens were collected. Pre- and post-intervention tissue biopsies and stool samples were snap frozen in liquid nitrogen and saved at −80°C. A set of colon mucosal biopsies were fixed in 10% buffered formalin. A venous blood sample was drawn for serum liver function and injury markers at both visits. For each of the two time-points, one biopsy and one stool specimen from each participant was utilized for microbiome analysis and one biopsy and one stool specimen was utilized for metabolomic profiling. The remaining tissue samples were used for histological, immunohistochemical and proteomic analyses. Microbiome / metabolomic data from this trial have been published along with safety and tolerability findings [28]. This report presents findings from histology / immunohistology and initial proteomic analysis.

During the participation phase, study coordinators remotely contacted subjects monthly to assess adherence to the study protocol and to identify any adverse events. Study compliance was evaluated by counting unused capsules and capsule log entries returned at the end of the study. These findings were presented in a recent report [28].

### Histology and quantitative immunohistochemistry

Baseline and post intervention colon mucosal biopsies were fixed in 10% buffered formalin and embedded in paraffin. Paraffin-embedded biopsies were sectioned and stained with hematoxylin and eosin (H&E) and examined by light microscopy for tissue orientation and full-length crypt evaluation.

For immunohistochemistry (IHC), paraffin-embedded tissue was sectioned and assessed for markers for proliferation (Ki67) and differentiation (Cytokeratin20 [CK20] and p21 [or WAF1]). Supplement Table 1 provides a list of antibodies used, their source and relevant characteristics. These antibodies have been tested and validated for the formalin-fixed, paraffin-embedded tissue. Briefly, colonic samples were sectioned at 5 μm thickness, deparaffinized, rehydrated in graded ethanol solutions and subjected to heat-induced epitope retrieval with high pH or low pH FLEX TRS Retrieval buffer (Agilent Technologies, 154 #K8004; Santa Clara, CA, United States) for 20 min. After peroxidase blocking, antibodies were applied at appropriate dilutions at room temperature for 30 or 60 min depending on the manufacturer’s recommendation. The FLEX HRP EnVision System (Agilent Technologies) was used for detection with a 10-min DAB chromagen application. Control tissue was used to test for negative and positive staining.

### Quantitative morphometry

Colonic sections of stained and immunostained tissue on glass slides were digitally scanned (at 40X) using the Aperio AT2 brightfield whole slide scanner (Leica Biosystems) at a resolution of 0.5 µm per pixel with 20X objective for quantitative image analysis. The scanned images were housed on a secure server and remotely accessed using Leica Aperio eSlide Manager (v12.4.3.5008), a digital pathology management software. A slide-viewing software by Aperio ImageScope (v12.4.3.5008) and associated image analysis tools were used to examine and quantitate these scanned histological sections.

For crypt length measurement, digitized H&E stained sections were analyzed to evaluate full-length crypts by measuring the height of each crypt from the base of the crypt to the mucosal surface. Crypt lengths were averaged for each slide, grouped and compared with the other groups. Brightfield Immunohistochemistry Image Analysis tools (Leica) were used to quantify and interpret biomarker expression of the three immunostains used in this study. Crypts were selected and lamina propria was excluded using annotation tool. Markup images were checked for the accuracy of the quantitation. Aperio Nuclear Algorithm (v9) was used for Ki67 and p21 expression quantification. This algorithm measures intensity of the nuclear staining in individual cells and separates those into very intense to no nuclear staining (3+, 2+, 1+, and 0 respectively). Percent positive nuclei within the crypts were used here for comparison. Nuclei (of cells) in the lamina propria and submucosa were not included. To quantify CK20 expression, the Aperio Positive Pixel Count Algorithm (v9) was used. It quantifies the number and intensity of pixels of a specific stain in a digitized image. Positivity was calculated with numbers of positive pixels against total pixels present within the colonic crypts and mucosal surface.

### Proteomic assessment

Proteomic experiments were performed at the Proteomics Resource Facility (PRF) in the Department of Pathology at the University of Michigan, employing mass spectrometry (MS)-based tandem mass tag (TMT) analysis (ThermoFisher Scientific) for the relative quantification of proteins. Individual colon biopsies were weighed (the weights were ranged from 5.5 mg to 12 mg) and exposed to Radioimmunoprecipitation Assay (RIPA) Lysis and Extraction buffer (Thermo Scientific cat#89901) at 25 µl per mg for protein. After that, biopsies were manually homogenized employing 1.5 ml tube sample pestles (RPI item#199228) and then freeze fractured for 1 cycle. Samples were centrifuged at 20,000 g for 10 min to remove insoluble material. Supernatants were removed to a clean tube and then protein concentrations were determined using a BCA assay kit (Pierce cat#23227). Following this, individual samples were “pooled” in groups of five (i.e., 5 placebo, 5 calcium and 5 Aquamin^®^ specimens, before and after intervention), allowing us to evaluate all 30 samples in two runs. Each pool was comprised of 8 micrograms per sample, 5 samples per pool for a total of 40 µg. The volume was adjusted such that each pool was 40 µg of pooled protein in 40 µl total volume. Briefly, forty micrograms of protein from each pool were separately digested with trypsin and individually labeled with one of the 6 isobaric mass tags following the manufacturer’s protocol using TMTsixplex kit (ThermoFisher cat# 90061) for each MS analysis. After labeling, equal amounts of the peptide from each pool were mixed together. In order to achieve in-depth characterization of the proteome, the labeled peptides were fractionated using 2D-LC (basic pH reverse-phase separation followed by acidic pH reverse phase) and analyzed on a high-resolution, tribrid mass spectrometer (Orbitrap Fusion Tribrid, ThermoFisher Scientific) using conditions optimized at the PRF. MultiNotch MS3 approach [32] was employed to obtain accurate quantitation of the identified proteins/peptides. Data analysis was performed using Proteome Discoverer (v2.4, ThermoFisher). MS2 spectra were searched against UniProt human protein database (downloaded on 06/20/2019; 20353 reviewed entries) using the following search parameters: MS1 and MS2 tolerance were set to 10 ppm and 0.6 Da, respectively; carbamidomethylation of cysteines (57.02146 Da) and TMT labeling of lysine and N-termini of peptides (229.16293 Da) were considered static modifications; oxidation of methionine (15.9949 Da) and deamidation of asparagine and glutamine (0.98401 Da) were considered variable. Identified proteins and peptides were filtered to retain only those that passed ≤2% false-discovery rate (FDR) threshold of detection. Quantitation was performed using high-quality MS3 spectra (average signal-to-noise ratio of 6 and 75% isolation interference). Protein names were retrieved using Uniprot.org, and Reactome v74 (reactome.org) was used for initial pathway enrichment analysis. STRING database - v11 (string-db.org) was used to detect protein-protein interactions and additional enrichment analyses provide information related to cellular components, molecular functions and biological processes by Gene Ontology (GO) annotation. It also offered Reactome and Kyoto Encyclopedia of Genes and Genomes (KEGG) databases to curate pathways. Only proteins with a ≤2% FDR confidence were included in the analyses. The differential protein expression profiles were established by calculating the abundance ratios of normalized abundances of post-intervention samples to pre-intervention samples. For the final comparison, the pre-intervention samples from all 30 subjects and the post-intervention samples from the placebo group were combined. Final abundance ratios of post-intervention samples were compared to this. It should be noted that, when only the respective pre-treatment values for each cohort were used as control or when the pre-treatment values from the three cohorts were combined (without the post-treatment placebo) were used to assess proteomic expression, the findings were similar.

The initial analysis involved an unbiased proteome-wide screen of all proteins modified by Aquamin^®^ or by calcium alone in relation to placebo. Follow-up analysis was targeted toward proliferation, differentiation and barrier-related cell adhesion proteins. Mass spectrometry based proteomics data were deposited to the ProteomeXchange Consortium via the PRIDE partner repository and the identifier is pending.

### Data analysis and statistical evaluation

Pre- post-intervention ordinates were obtained for each discrete morphological and immunohistochemical feature. Means and standard deviations were generated, and data were analyzed by ANOVA followed by the two-stage linear step-up procedure of Benjamini, Krieger and Yekutieli for multiple comparison (by controlling FDR). For correlation between two markers, Pearson correlation coefficient was computed at 95% confidence interval with two-tailed p value ≤0.05. GraphPad Prism v8.3 was used for these analyses. Proteomic data were generated in two separate runs of six pooled samples per run. Data from the two runs were combined. Results from subjects treated with Aquamin^®^ or calcium alone were compared to results obtained with the placebo. Pathways enrichment data reflect Reactome-generated p-values based on the number of entities identified in a given pathway compared to total proteins involved in that pathway. For STRING enrichment analysis, the whole genome statistical background was assumed and FDR stringency was high (≤1%). Data were considered significant at p and q ≤0.05.

## RESULTS

### Histological and immunohistological findings: Comparison of Aquamin^®^ with calcium alone and placebo

In the first series of studies, colonic biopsies were examined at the light microscopic level for overall crypt appearance and assessed morphometrically for crypt length. Crypt length was chosen since previous studies have demonstrated little change in this parameter with calcium intervention [31]. Crypt length measurement data from all 30 subjects (10 in each group) are presented in Figure 1A and Supplement Figure 1A. As can be seen from the Figure 1A, there were no apparent effects of the three interventions on crypt length.

**Figure 1.**
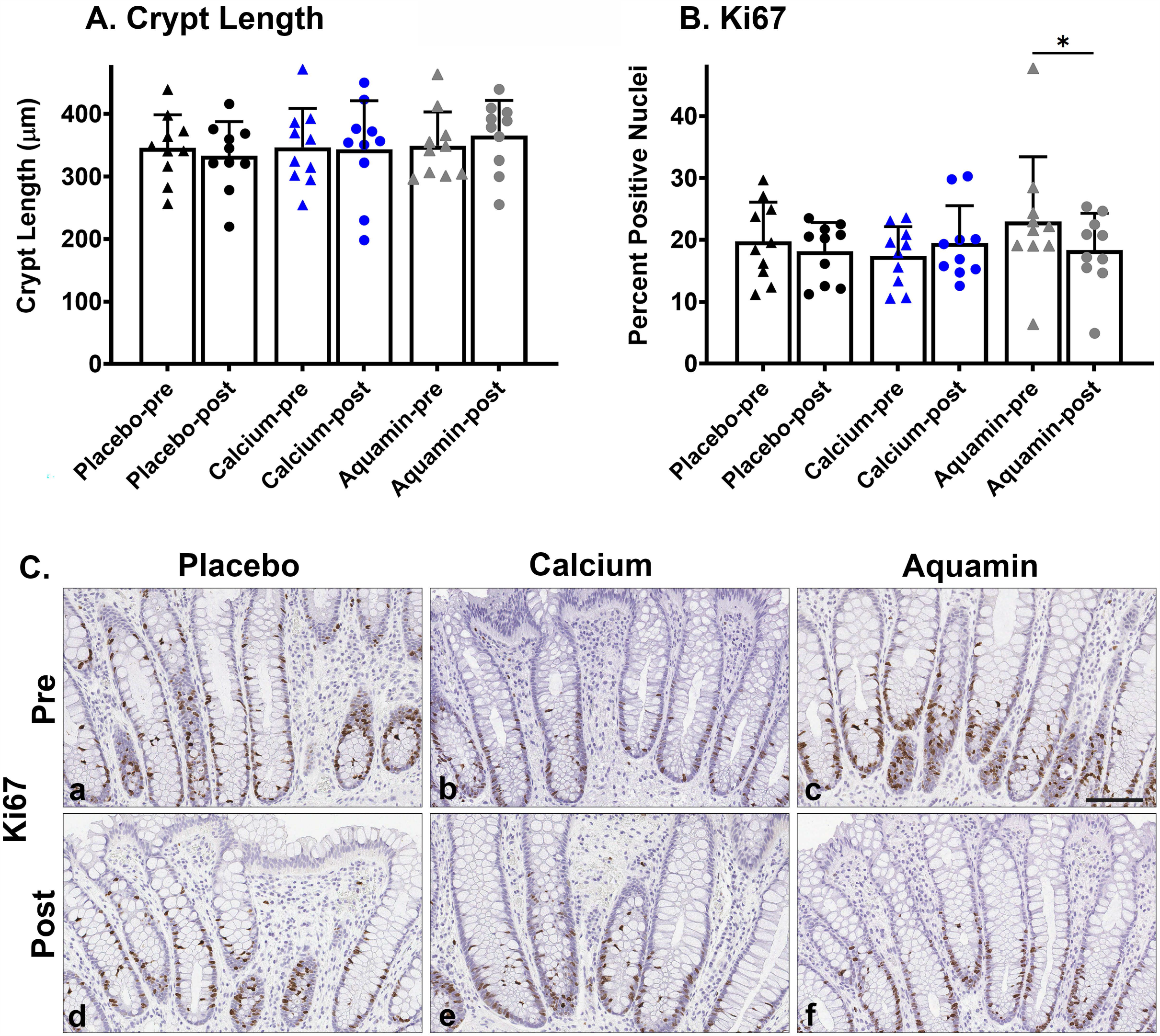
Histological features of the colonic mucosa and proliferation expression. **A:** Crypt Length. Values represent means and standard deviations based on measurement of crypt length in individual crypts (P:70 & 85, C:2 & 71, AQ:74 & 96) per treatment group (10 subjects per group before and after treatment). **B:** Ki67 expression quantitation. Percentage of Ki67-positive nuclei is presented in each group. Values represent means and standard deviations based on assessment of individual crypts (P=106 & 81, C=63 & 65, AQ=102 & 82) per treatment group (10 subjects per group before and after 90-day intervention). *indicates significance as compared to the respective pre-intervention value if analysis is done based on the number of crypts in each group (p and q ≤0.0001). **C:** Histological appearance and Ki67-stained histological images of the colonic mucosa from a representative subject in each treatment group before and after the 90-day intervention. Scale bar = 100 µm.

Results (quantitative data) of immunostaining studies with Ki67 are shown in Figure 1B. Images of representative sections of pre- and post-treatment biopsies from one subject in each cohort are presented in Figure 1C. Values from the placebo group and calcium group did not differ significantly between pre-treatment and post-treatment. In the Aquamin^®^-treated cohort, however, there was a modest reduction (approximately 20%) in the average post-treatment value compared to average pre-treatment value (p and q ≤0.05 when the data was analyzed using the total number of crypts in each treatment group). As expected, virtually all of the Ki67 staining was in cells occupying the lower third of the crypt. The complete data set (from each individual) is presented in Supplement Figure 1B.

Figure 2 presents immunostaining results with CK20 and p21. With CK20, no change in expression was observed with either intervention. In both pre- and post-intervention specimens, there was strong brown staining visible on the luminal mucosal surface and the top one-third of the full-length crypts (Figure 2A). With p21, there was a trend toward increased expression with both calcium alone (by 117%) and Aquamin^®^ (by 99%) in post-intervention biopsies as compared to baseline expression (p and q ≤0.05 when the data were analyzed using the total number of crypts in each treatment group). Positive nuclear staining with p21 was visible in cells on the mucosal surface and the top half of colonic crypts (Figure 2B). The trend toward increased p21 expression in response to calcium is consistent with previous findings [29, 30]. The pre-post data from each individual are presented in Supplement Figure 1C and 1D.

**Figure 2.**
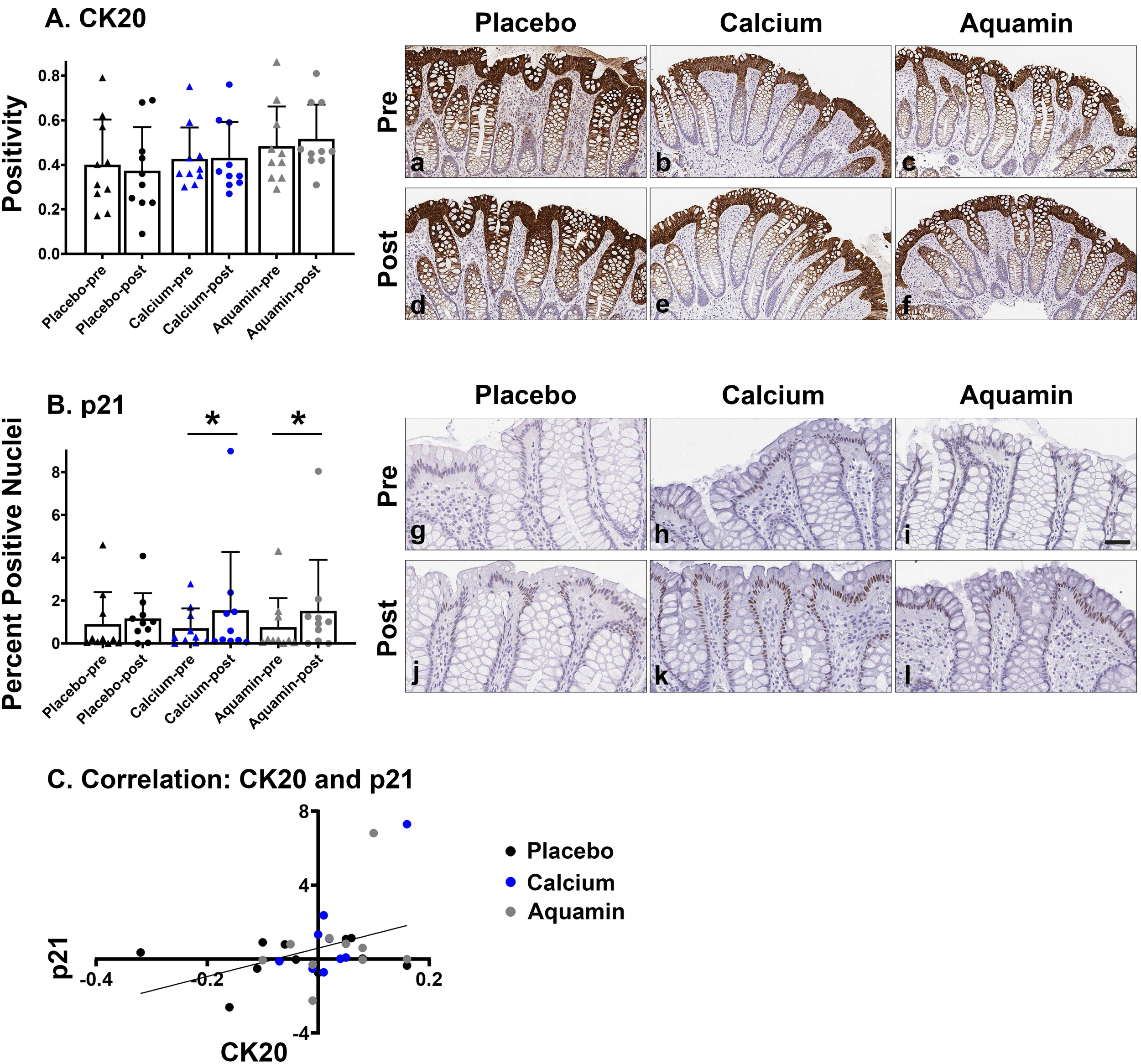
CK20 and p21 expression. **A**: CK20 expression quantitation. CK20 expression is presented by CK20 stain positivity. Values represent means and standard deviations based on assessment of individual crypts (P:106 & 81, C:63 & 65, AQ:102 & 82) and luminal surface per treatment group (10 subjects per group before and after treatment for 90 days). Inset (a-f): CK20-stained images of colonic mucosa from a representative subject of each treatment group before and after the 90-day intervention. Scale bar = 100 µm. **B:** p21 expression quantitation. Percentage of strong positive (2+ and 3+) nuclei is used to present p21 expression. Values represent means and standard deviations based on assessment of individual crypts (P:106 & 78, C:59 & 63, AQ:103 & 65) and luminal epithelial cells per treatment group (10 subjects per group before and after treatment for 90 days). *indicates significance as compared to the respective pre-intervention value if analysis is done based on the number of crypts in each group (C: p=0.0084 and q=0.0044; AQ: p=0.0058 and q=0.0044). Inset (g-l): p21-stained images of colonic mucosa from a representative subject of each treatment group before and after the 90-day intervention. Scale bar = 50 µm. **C:** Correlation of CK20 and p21 expressions in all 30 subjects. r = 0.3819; p (two-tailed) =0.0373

As part of the immunohistochemical analyses, we searched for correlations between marker expression patterns. Neither CK20 nor p21 expression demonstrated a (positive or negative) correlation with Ki67 expression but the two differentiation markers were positively correlated (p= 0.0373) with each other (Figure 2C).

### Proteomic findings: Unbiased comparison of Aquamin^®^ with calcium alone and placebo

Each of the three interventions was assessed for effects on protein expression profile as described in the Materials and Methods section. A total of 5847 distinct proteins appeared in the mass spectrometry (MS)-based TMT analysis with FDR of ≤2%. An overview of the initial unbiased proteomic screen is presented in Figure 3. The pooled data from all three pre-treatment groups and the post-placebo treatment group were used as a control to give the most comprehensive view of the protein-expression changes in response to each intervention. The data presented in Figure 3A were calculated at 1.5-fold change. The bar graphs on the top left represent the total number of altered proteins at 1.5-fold with ≤2% FDR for each of the three interventions. On the right, two Venn diagrams are presented to show the overlap in altered proteomic expression among the three interventions. It is evident from these data that most of the protein alterations (either up- or downregulated) occurred in response to Aquamin^®^ or calcium alone; there was minimal change in the placebo control group. Similarly, when the stringency was lenient (1.1-fold), the overall trend was the same; that is, most of the alteration was seen with Aquamin^®^ or calcium (Figure 3B). Aquamin^®^ still showed the highest change in quantitative proteomic expression. A heatmap was used to present the differences in proteomic expression among three interventions at 1.5 fold-change, upregulated in Figure 3C and downregulated in Figure 3D. Our conclusion from the unbiased analysis is that the 90-day period of treatment with either Aquamin^®^ or calcium alone was sufficient to detect an effect on the protein expression pattern in the colonic mucosa of healthy adult volunteers that was not observed with placebo.

**Figure 3.**
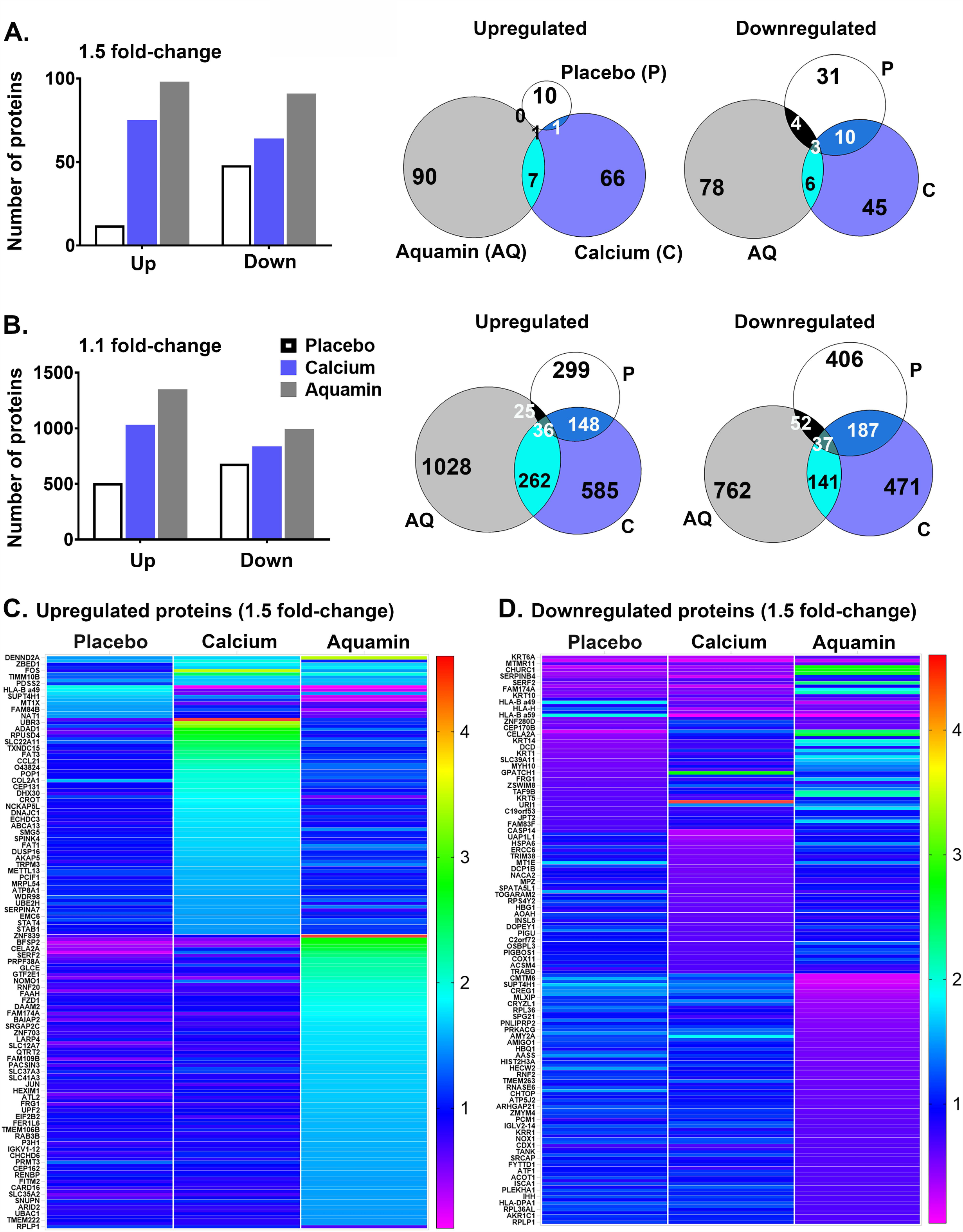
Proteomic landscape of colon mucosal biopsies and response to interventions: **A**. Upregulated and downregulated proteins in each cohort at 1.5-fold. Each bar represents the number of proteins up-regulated (1.5-fold; ≤2% FDR) or down-regulated (0.67-fold; ≤2% FDR) over the 90-day course of treatment with each of the three interventions. On the right, Venn diagrams show the overlap of these upregulated and downregulated moieties to provide common proteins among 3 groups or between 2 groups or unique to an intervention. The list of these upregulated proteins is presented in Supplement Table 2. While downregulated proteins are presented in Supplement Table 3. **B**. Upregulated and downregulated proteins in each cohort at 1.1-fold. Each bar represents the number of proteins up-regulated (1.1-fold; ≤2% FDR) or down-regulated (0.9-fold; ≤2% FDR) over the 90-day course of treatment with each of the three interventions. On the right, Venn diagrams show the overlap of these upregulated and downregulated moieties to provide common proteins among 3 groups or between 2 groups or unique to an intervention. **C**: Differential proteomic expression of upregulated proteins (of all interventions at 1.5-fold) is presented in a heatmap. **D**: Differential proteomic expression of downregulated proteins (of all interventions at 1.5-fold) is presented in a heatmap.

As part of the analysis, we identified individual proteins responsible for altered expression at 1.5-fold change. The individual proteins (assessed by unbiased screen) are presented in Supplement Table 2 (up-regulated) and in Supplement Table 3 (down regulated). The proteins presented in Supplement Tables 2 and 3 were sorted based on the overlap among all three interventions or between two interventions or unique to each intervention. As a next step, we used STRING enriched analysis to highlight any protein-protein interactions among proteins presented in Supplement Tables 2 and 3. Some clusters of proteins were evident by having multiple interactions among the participating proteins (Supplement Figure 2).

Similarly, Supplement Table 4 lists all the STRING-enriched GO biological processes, molecular functions and cellular components significantly (with q ≤0.05) altered with the same set of unbiased proteins. In the final step, significantly altered pathways (with q ≤0.05) were identified (by Reactome and KEGG curated in STRING) using unbiased proteins altered at 1.5-fold (shown in Supplement Figure 3). The top pathways involved by the altered proteins were following; keratinization, cornified envelope formation, hemidesmosome assembly and collagen related pathways were up and pathways related to mucosal inflammation, antigen processing / presentation, and viral carcinogenesis were down (Supplement Figure 3).

### Proteomic findings: Directed search

We next searched the database for differentiation-related proteins (keratins) along with proteins involved in cell-cell and cell-matrix adhesion (adherens junction, tight junction and desmosomal proteins) and other moieties that contribute to barrier structure / function in the colon. These include basement membrane components, carcinoembryonic antigen-related cell adhesion molecules (CEACAMs), mucins, Nectin1, a cell adhesion molecule, and collagen chains. As can be seen from the Figure 4, substantial up-regulation (compared to placebo) was observed with many of these proteins in biopsies from subjects ingesting either Aquamin^®^ or calcium alone. For the majority of proteins shown in the Figure 4, up-regulation with Aquamin^®^ was greater than with calcium alone. Most of the identified moieties are epithelial cell products. However, treatment with Aquamin^®^ and / or calcium alone also up-regulated the expression of several collagen chains. Altered expression of collagen chains was not prominent in our earlier colonoid studies [23, 24] but seen here; reflecting the fact that the colon biopsies have both epithelial and stromal components. Certain collagen chains (Collagen IV) are integral to basement membrane and others mediate cell attachment to basement membrane [33, 34]. The Reactome database was used to identify pathways affected by the proteins up-regulated in response to Aquamin^®^ or calcium alone (Table 1). Not surprisingly, pathways involved in cellular adhesive functions, laminin interactions and extracellular matrix organization were among the most highly affected.

**Table 1.** Significantly Altered Pathways (by upregulated proteins with targeted approach)

| **Pathway name** | **Entities pValue** | **Entities FDR** | **Mapped entities** |
| --- | --- | --- | --- |
| Laminin interactions | 1.1x10<sup>-16</sup> | 1.9x10<sup>-14</sup> | COL4A2;HSPG2;LAMA4;LAMA5;LAMB1;LAMB2;LAMB3;LAMC1;NID2 |
| ECM proteoglycans | 3.0x10<sup>-15</sup> | 2.3x10<sup>-13</sup> | COL4A2;COL6A1;COL6A2;FN1;HSPG2;LAMA4;LAMA5;LAMB1;LAMB2;LAMC1 |
| Extracellular matrix organization | 4.1x10<sup>-15</sup> | 2.3x10<sup>-13</sup> | CEACAM1;COL23A1;COL4A2;COL6A1;COL6A2;FN1;HSPG2;LAMA4;LAMA5;LAMB1;LAMB2;LAMB3;LAMC1;NID2 |
| Non-integrin membrane-ECM interactions | 2.4x10<sup>-14</sup> | 9.9x10<sup>-13</sup> | COL4A2;FN1;HSPG2;LAMA4;LAMA5;LAMB1;LAMB2;LAMB3;LAMC1 |
| Degradation of the extracellular matrix | 1.2x10<sup>-12</sup> | 3.6x10<sup>-11</sup> | COL23A1;COL4A2;COL6A1;COL6A2;FN1;HSPG2;LAMA5;LAMB1;LAMB3;LAMC1 |
| MET activates PTK2 signaling | 1.3x10<sup>-12</sup> | 3.6x10<sup>-11</sup> | FN1;LAMA4;LAMA5;LAMB1;LAMB2;LAMB3;LAMC1 |
| MET promotes cell motility | 1.1x10<sup>-11</sup> | 2.7x10<sup>-10</sup> | FN1;LAMA4;LAMA5;LAMB1;LAMB2;LAMB3;LAMC1 |
| Formation of the cornified envelope | 9.1x10<sup>-10</sup> | 1.9x10<sup>-08</sup> | DSP;KRT1;KRT10;KRT14;KRT20;KRT6A;KRT9; KRT77 |
| Signaling by MET | 1.1x10<sup>-09</sup> | 2.0x10<sup>-08</sup> | FN1;LAMA4;LAMA5;LAMB1;LAMB2;LAMB3;LAMC1; |
| Keratinization | 5.0x10<sup>-08</sup> | 8.1x10<sup>-07</sup> | DSP;KRT1;KRT10;KRT14;KRT20;KRT6A;KRT9;KRT77 |
| Integrin cell surface interactions | 6.9x10<sup>-08</sup> | 1.0x10<sup>-06</sup> | COL23A1;COL4A2;COL6A1;COL6A2;FN1;HSPG2 |
| Signaling by Receptor Tyrosine Kinases | 3.6x10<sup>-07</sup> | 5.0x10<sup>-06</sup> | COL4A2;COL6A1;COL6A2;FN1;LAMA4;LAMA5;LAMB1;LAMB2;LAMB3;LAMC1 |
| Developmental Biology | 1.0x10<sup>-06</sup> | 1.2x10<sup>-05</sup> | COL4A2;COL6A1;COL6A2;DSP;KRT1;KRT10;KRT14;KRT20;KRT6A;KRT9;KRT77;LAMB1;LAMC1 |
| Collagen formation | 3.1x10<sup>-06</sup> | 3.7x10<sup>-05</sup> | COL23A1;COL4A2;COL6A1;COL6A2;LAMB3 |
| Collagen chain trimerization | 4.8x10<sup>-06</sup> | 5.3x10<sup>-04</sup> | COL23A1;COL4A2;COL6A1;COL6A2 |
| Assembly of collagen fibrils and other multimeric structures | 1.7x10<sup>-05</sup> | 1.7x10<sup>-04</sup> | COL4A2;COL6A1;COL6A2;LAMB3 |
| Collagen degradation | 2.1x10<sup>-05</sup> | 1.9x10<sup>-04</sup> | COL23A1;COL4A2;COL6A1;COL6A2 |
| Collagen biosynthesis and modifying enzymes | 2.5x10<sup>-05</sup> | 2.2x10<sup>-04</sup> | COL23A1;COL4A2;COL6A1;COL6A2 |
| Cell junction organization | 8.5x10<sup>-05</sup> | 0.001 | CLDN3;KRT14;LAMB3;NECTIN1 |
| Fibronectin matrix formation | 1.1x10<sup>-04</sup> | 0.001 | CEACAM1;FN1 |
| Post-translational protein phosphorylation | 1.5x10<sup>-04</sup> | 0.001 | FN1;LAMB1;LAMB2;LAMC1 |
| NCAM1 interactions | 1.7x10<sup>-04</sup> | 0.001 | COL4A2;COL6A1;COL6A2 |
| Regulation of IGF Factor transport and uptake by IGFBPs | 2.6x10<sup>-04</sup> | 0.002 | FN1;LAMB1;LAMB2;LAMC1 |
| Cell-Cell communication | 3.2x10<sup>-04</sup> | 0.002 | CLDN3;KRT14;LAMB3;NECTIN1 |
| Type I hemidesmosome assembly | 3.7x10<sup>-04</sup> | 0.002 | KRT14;LAMB3 |
| Apoptotic cleavage of cell adhesion proteins | 3.7x10<sup>-04</sup> | 0.002 | DSP;OCLN |
| Signaling by PDGF | 4.8x10<sup>-04</sup> | 0.003 | COL4A2;COL6A1;COL6A2 |
| NCAM signaling for neurite out-growth | 5.7x10<sup>-04</sup> | 0.003 | COL4A2;COL6A1;COL6A2 |
| Anchoring fibril formation | 6.8x10<sup>-04</sup> | 0.003 | COL4A2;LAMB3 |
| Defective GALNT12 causes colorectal cancer 1 | 0.001 | 0.005 | MUC2;MUC4 |
| Termination of O-glycan biosynthesis | 0.002 | 0.01 | MUC2;MUC4 |
| Apoptotic cleavage of cellular proteins | 0.004 | 0.02 | DSP;OCLN |
| Apoptotic execution phase | 0.01 | 0.03 | DSP;OCLN |
| Dectin-2 family | 0.01 | 0.04 | MUC2;MUC4 |
| O-linked glycosylation of mucins | 0.01 | 0.04 | MUC2;MUC4 |
| Cell-cell junction organization | 0.01 | 0.04 | CLDN3;NECTIN1 |
| RUNX1 regulates expression of components of tight junctions | 0.01 | 0.04 | OCLN |
| PTK6 Regulates Cell Cycle | 0.02 | 0.05 | CDKN1B |
| Nectin/Necl trans heterodimerization | 0.02 | 0.05 | NECTIN1 |

Insulin-like Growth Factor: IGF; Insulin-like Growth Factor Binding Protein: IGFBP; Neural Cell Adhesion Molecule: NCAM.

**Figure 4.**
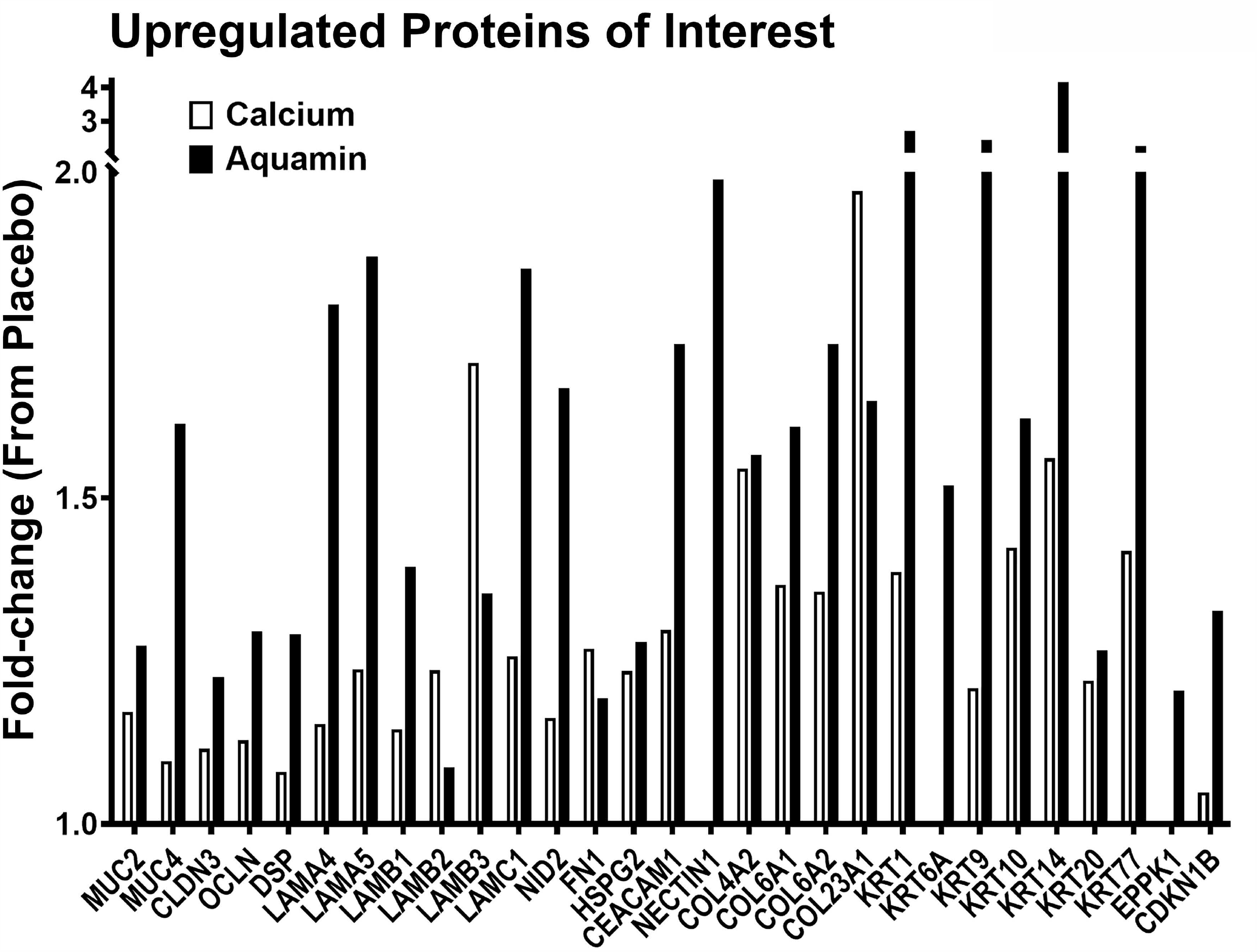
Upregulated proteins of interest. Fold-change value of each protein is presented in response to placebo.

The database was also searched for proteins involved in proliferation, DNA synthesis, tumor metastasis, Wnt/β-catenin signaling and inflammation. The overexpression of some of these proteins is associated with certain cancers. Several such moieties were identified. Most of these were down-regulated (Figure 5) with Aquamin^®^. Interesting, many of the growth-related proteins that were decreased in response to Aquamin^®^ were not substantially altered with calcium alone. Supplement File 1 provide relevance and further information (including citations) related to the proteins in Figure 5.

**Figure 5.**
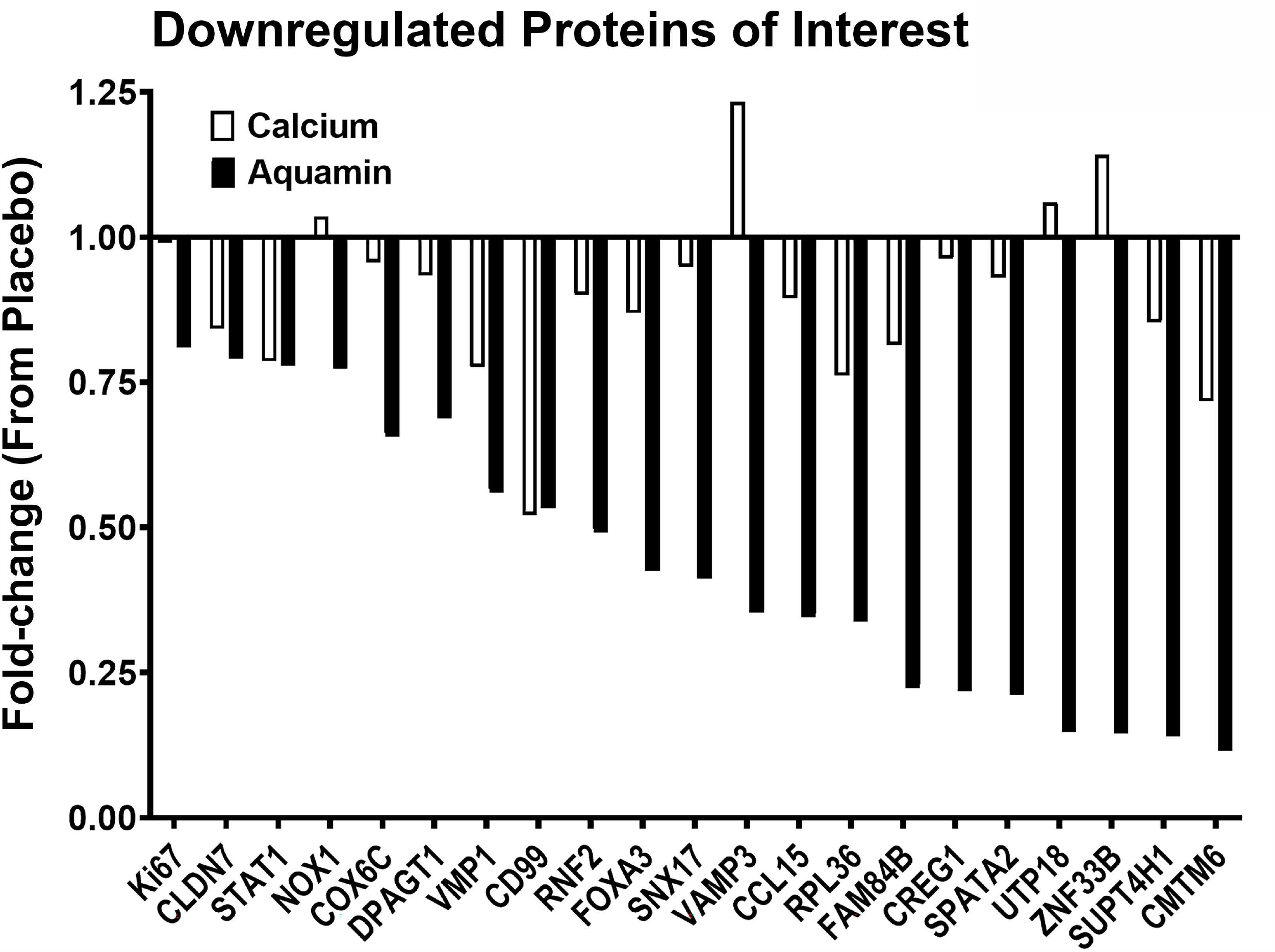
Downregulated proteins of interest. Fold-change value of each protein is presented in response to placebo.

## DISCUSSION

We recently completed a 90-day interventional trial in which Aquamin^®^, a calcium- and magnesium-rich multi-mineral natural product derived from red marine algae, was shown to be well-tolerated by healthy adult individuals [28]. The same study demonstrated potentially beneficial effects of Aquamin^®^ based on alterations in gut microbial composition and attendant metabolomic profile. The microbial and metabolomic effects were not observed in a placebo-treated cohort or in subjects treated with calcium alone at an equivalent amount to that provided with Aquamin^®^. Additional findings from the same interventional trial are reported in the current manuscript. Here we show that while histological features (overall colonic crypt appearance and crypt length) and a immunohistochemical marker of differentiation (CK20) demonstrated no change in biopsies from Aquamin^®^-treated subjects relative to those receiving placebo. Meanwhile a marker of proliferation (Ki67) was reduced modestly with Aquamin^®^ treatment and a marker of cell differentiation (p21) was increased. Consistent with findings of others, calcium alone did not induce a measurable decrease in Ki67 expression [30], but induced increased expression of p21 [29, 30].

In parallel with the histological / immunohistological findings, we utilized a proteomic approach to assess the effects of Aquamin^®^ or calcium alone on protein profile in the colonic mucosa. The data generated in the proteomic screen showed that the 90-day treatment was sufficient to demonstrate a response. Regardless of whether we used only the respective pre-treatment values from each cohort (n=10 per group) as control, or merged all of the pre-treatment values (n=30 per group) or merged the pretreatment values and the post-treatment values from the placebo cohort (n=40 per group) to generate a control, the numbers of proteins up- and down-regulated) was greater with Aquamin^®^ than the number affected in the placebo group. Among up-regulated proteins, there were several that define the differentiated state and others that contribute to cell-cell and cell-matrix adhesion functions (see Figures 3 and 4). Among down-regulated proteins was Ki67 and several other moieties that contribute to growth-regulation (Figure 5). Also, a protein similar to p21 known as CDKN1B or p27 was upregulated with Aquamin^®^. It is a regulator of cell cycle progression [35]. These findings are potentially significant from a number of standpoints, as discussed below.

The data presented here and in our recent report from the same trial [28] show that a 90-day course of daily treatment with Aquamin^®^ providing 800 mg of calcium per day is sufficient to obtain a measurable effect on markers of differentiation and proliferation in the colonic mucosa as well as to modulate gut microbiome / metabolomic parameters. The effects observed with Aquamin^®^ were not duplicated with calcium alone (though calcium alone did, in fact, alter expression of some of the same markers as Aquamin^®^). Consistent with these findings, our past long-term studies in rodents demonstrated multiple health benefits with Aquamin^®^ (reduced colon polyp incidence [26, 27], reduced liver injury [36], improved bone mineralization [37, 38] and reduced inflammatory skin disease [39]) that were only partially mimicked with calcium alone, while our studies with human colon tissue in organoid culture [22, 23] also provided evidence for beneficial activity with Aquamin^®^ not seen with calcium alone. Together, these observations support continuation of efforts to demonstrate the utility of a multi-mineral approach for chronic disease prevention and continuation of drug-development efforts with Aquamin^®^ itself.

Although the major focus of this effort was colon polyp prevention, our findings allow us to suggest that benefits of multi-mineral intervention may extend beyond this indication. Specifically, the up-regulation of multiple adhesion molecules, both in the interventional model described here and in organoid culture model [22-25], may translate into improved colon mucosal barrier. In the organoid culture model, it should be noted, exposure to Aquamin^®^ resulted in increased trans-epithelial electrical resistance and increased tissue cohesion in parallel with adhesion molecule and barrier protein up-regulation [25]. Defects in barrier function – allowing gut bacteria, food allergens and potentially toxic substances access to the interstitial space – are likely to contribute to inflammatory bowel diseases (IBD) including Crohn’s Disease and ulcerative colitis [40, 41]. A loss of permeability control has also been noted in relation to irritable bowel syndrome [42]. More recently, it has been suggested that chronic systemic inflammation associated with obesity occurs, in part at least, as a consequence of high fat diet-induced defects in gastrointestinal permeability control [41]. While separating cause and effect in these various conditions is not straight forward, an improvement in barrier function would seem to be worthwhile in any case. Similarly, optimal gut barrier improvement may protect against CRC as well as inflammatory diseases [43]. Improved barrier function with a low-cost, low (to no)-toxicity agent could provide a cost-effective adjuvant therapy for a large group of individuals who suffer from chronic (and painful) inflammatory bowel conditions associated with barrier defects. Whether this will work, ultimately, and to what extent needs to be established experimentally in a carefully controlled clinical trial. To this end, we have recently initiated a pilot-phase trial (Clintrials.gov; NCT03869905), evaluating the efficacy of Aquamin^®^ as an adjuvant therapy for individuals with ulcerative colitis in remission. Results from the pilot-phase study along with our organoid culture data [23, 24, 28] and the results presented here should together provide an indication as to whether a multi-mineral approach to barrier improvement in the colon is helpful.

In addition to these upregulated proteins, the proteomic screen uncovered several interesting moieties that were downregulated with Aquamin^®^ (shown in Figure 5 and discussed in Supplement File 1). With several of these, overexpression plays a role in proliferation, cell cycle progression, chromatin organization, energy metabolism, and inflammation as well as tumor invasion and metastasis. For example, the knockdown of a protein known as E3 ubiquitin-protein ligase RING2 (RNF2) will inhibit cell proliferation and elevate p21 levels [44]. Similarly, CD99 antigen (CD99) is a cell adhesion molecule and involved in leukocyte transendothelial migration. It is correlated positively with active inflammatory bowel disease (IBD) disease activity [45]. CD99 is also expressed by tumor cells and have a role in tumor progression and cancer cell transendothelial migration [46]. CD99 was also downregulated in our previous study with UC-derived colonoid culture [24]. Another inflammation-related moiety, C-C motif chemokine 15 (CCL15) is a chemotactic factor that attracts T-cells and monocytes and is involved in inflammation. Loss of SMAD4 promotes CCL15 expression in CRC cells and enables primary tumor invasion and liver metastasis of CRC [47]. Interestingly, SMAD4 was upregulated with Aquamin^®^ (1.25 fold) while CCL15 was downregulated (by 0.35 fold). Together, these findings warrant additional studies to investigate the role of Aquamin^®^ in prevention of CRC in long-term polyp prevention trials.

The findings presented here, of course, must be taken as preliminary since the number of subjects (10 per group) was small and the treatment period (90 days) short. Due to these limitations, several (though not all) of the potentially important outcomes demonstrated beneficial trends but failed to reach a statistically significant level. Partially off-setting this limitation was the striking correlation between findings presented here and results from earlier studies in the colon organoid model [22-24]. In both the colon organoid model and the biopsies from the interventional trial (current study), we saw a modest reduction in the proliferation marker (Ki67) without a change in the differentiation marker (CK20) by immunohistology. Equally important, proteomic analysis revealed up-regulation of several cell-cell and cell-matrix adhesion molecules in both models, with basement membrane changes being especially prominent in both. Another limitation – one commonly faced in clinical trials – is the inability to collect functional response data to correlate with biomarker changes. Here again, the limitation was at least partially mitigated by our previous findings in organoid culture, where both TEER and cohesive function increased in Aquamin^®^-treated colonoids in parallel with adhesion molecule expression changes [25].

Another limitation: we simply do not know if the biomarker changes reported here with Aquamin^®^ will, ultimately, be reflective of a reduction in colon polyp incidence or with any other therapeutic benefit. The findings are sufficiently encouraging, however, to support further effort. As noted above, a therapeutic trial is currently under way. Equally important, these findings may be of value in so far as they could help other investigators by providing an indication of how large a change might be expected in several colonic biomarkers.

In summary, while it is well-known that having an adequate daily calcium intake is important for preventing various chronic, long-latency diseases [1-5], recent epidemiological studies have begun to appreciate the role of trace elements other than calcium in the mitigation of chronic diseases [12-15]. While our past preclinical studies [20-27] have provided compelling evidence that a multi-mineral approach with a natural product such as Aquamin^®^ can provide benefit not seen with calcium alone, results presented in our recent report [28] and those described here suggest that responses to multi-mineral intervention are robust enough to be seen in a small and comparatively simple clinical interventional trial setting.

## Data Availability

All the data are presented either as part of the manuscript or as supplemental files. The raw mass spectrometry proteomics data is submitted on ProteomeXchange Consortium (PRIDE partner repository), the identifier is pending.

## Acknowledgments

This study was supported by NIH grant CA201782 including supplemental funding through the Office of Dietary Supplements (to JV) and by an MCubed (University of Michigan) grant (to MNA & ILB). We thank Marigot LTD (Cork, Ireland) for providing Aquamin^®^ as a gift. We thank the Michigan Institute for Clinical and Health Research (MICHR), the Michigan Clinical Research Unit (MCRU), and Clinical Trials Support Office at the University of Michigan for their support with the trial. We thank Mr. Mitch Seymour from the MICHR IND/IDE Investigator Assistance Program (MIAP) for his help with IND process. We also thank Ms. Sherece Bank of Clinical Research Management (CRM) team at Michigan with her help with RedCap database. We also thank our study coordinators (Elaine Brady and Deepa Chandhrasekhar) for their assistance with the study. Most importantly, we are extremely thankful to the study participants who made it possible for us to conduct this study.

## FIGURE LEGENDS

**Supplement Figure 1. Change in histological and immunohistological features**. The pre-post difference in the colonic mucosa of each subject after 90 days of intervention. **A:** Crypt length. **B:** Ki67. **C:** CK20. **D:** p21.

**Supplement Figure 2. Unbiased proteomic screen: Protein interactions**. A network view of protein-protein interactions and predicted associations for the group of proteins at 1.5 fold-change (presented in Figure 3 and listed in Supplement Table 2 and Supplement Table 3) by employing STRING-database (v11). The strong interactions or associations can be seen with thick connecting lines. These lines are color-coded. Red line – indicates the presence of fusion evidence; Green line – neighborhood evidence; Blue line – co-occurrence evidence; Purple line – experimental evidence; Yellow line – text mining evidence; Light blue line – database evidence; Black line – co-expression evidence. There are multiple distinct clusters of proteins that are evident. The most prominent is in the placebo group with a set of downregulated proteins (Keratins & Cystatin-A [CSTA]).

**Supplement Figure 3. Unbiased proteomic screen: Altered pathways**. Most significant pathways with the altered proteins at 1.5 fold-change (presented in Figure 3 and listed in Supplement Table 2 and Supplement Table 3). String database (v11) was used for the enrichment analysis to generate altered pathways (KEGG and Reactome (v74) were the source databases for the pathways).

## Notes

### Competing Interest Statement

The authors have declared no competing interest.

### Clinical Trial

NCT02647671

### Clinical Protocols

https://clinicaltrials.gov/ct2/show/NCT02647671

### Author Declarations

This clinical interventional trial was conducted with FDA approval of Aquamin as an Investigational New Drug (IND#118194) and with oversight by the Institutional Review Board at the University of Michigan Medical School (IRBMED)-IRB#HUM00076276. The study was registered as an interventional clinical trial with details at Clinicaltrials.gov (study identifier NCT02647671). All participants provided written informed consent prior to inclusion. This phase I trial involving human participants was carried out in accordance with recognized ethical guidelines, for example, Declaration of Helsinki, International Ethical Guidelines for Biomedical Research Involving Human Subjects (CIOMS), the Belmont Report and the U.S. Common Rule.

