## Supplementary figures and images for "A MULTI-MINERAL INTERVENTION TO MODULATE COLONIC MUCOSAL PROTEIN PROFILE: Results from a 90-day trial in healthy human subjects"

### Supplement Figure 1

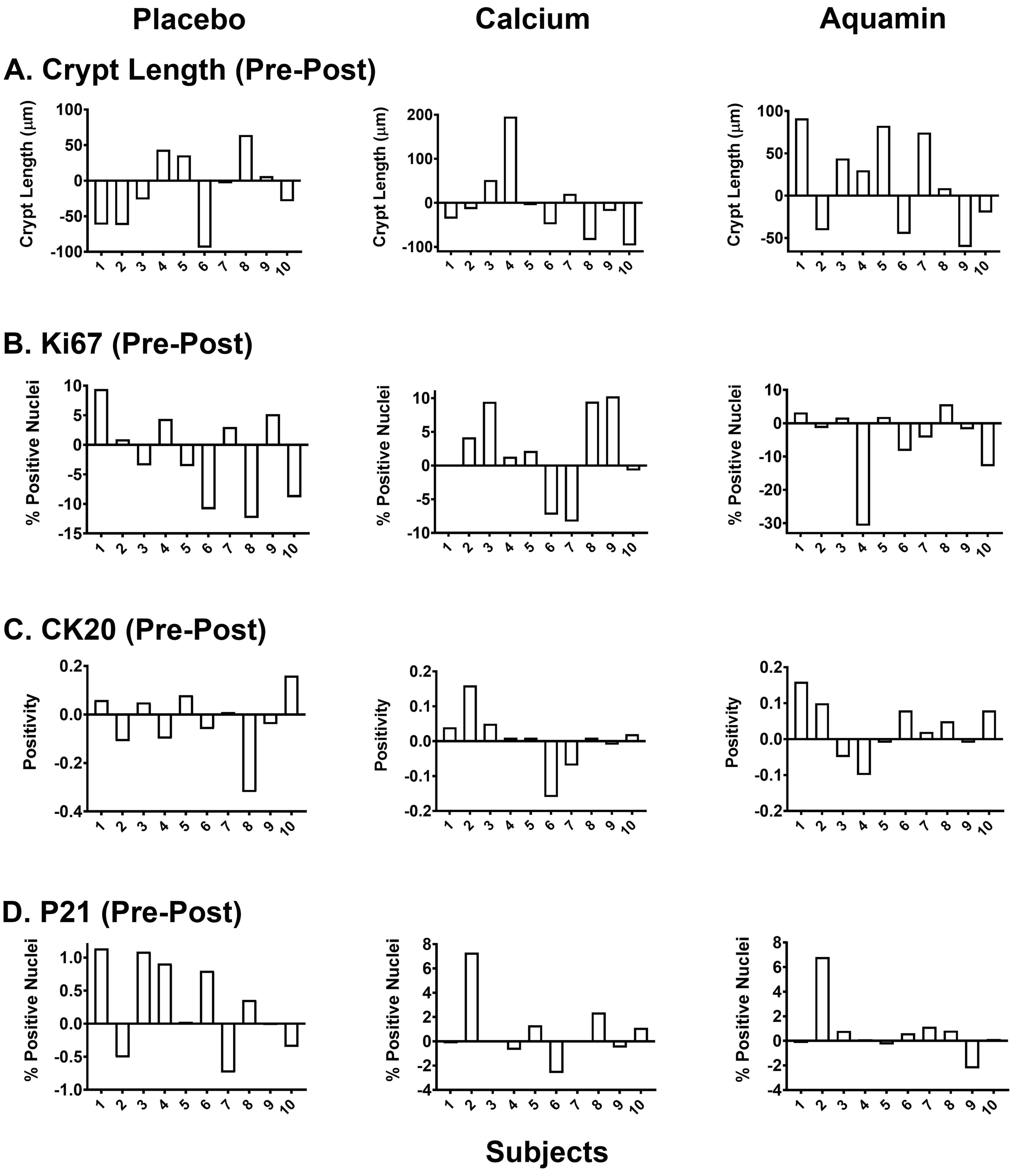

### Supplement Figure 2

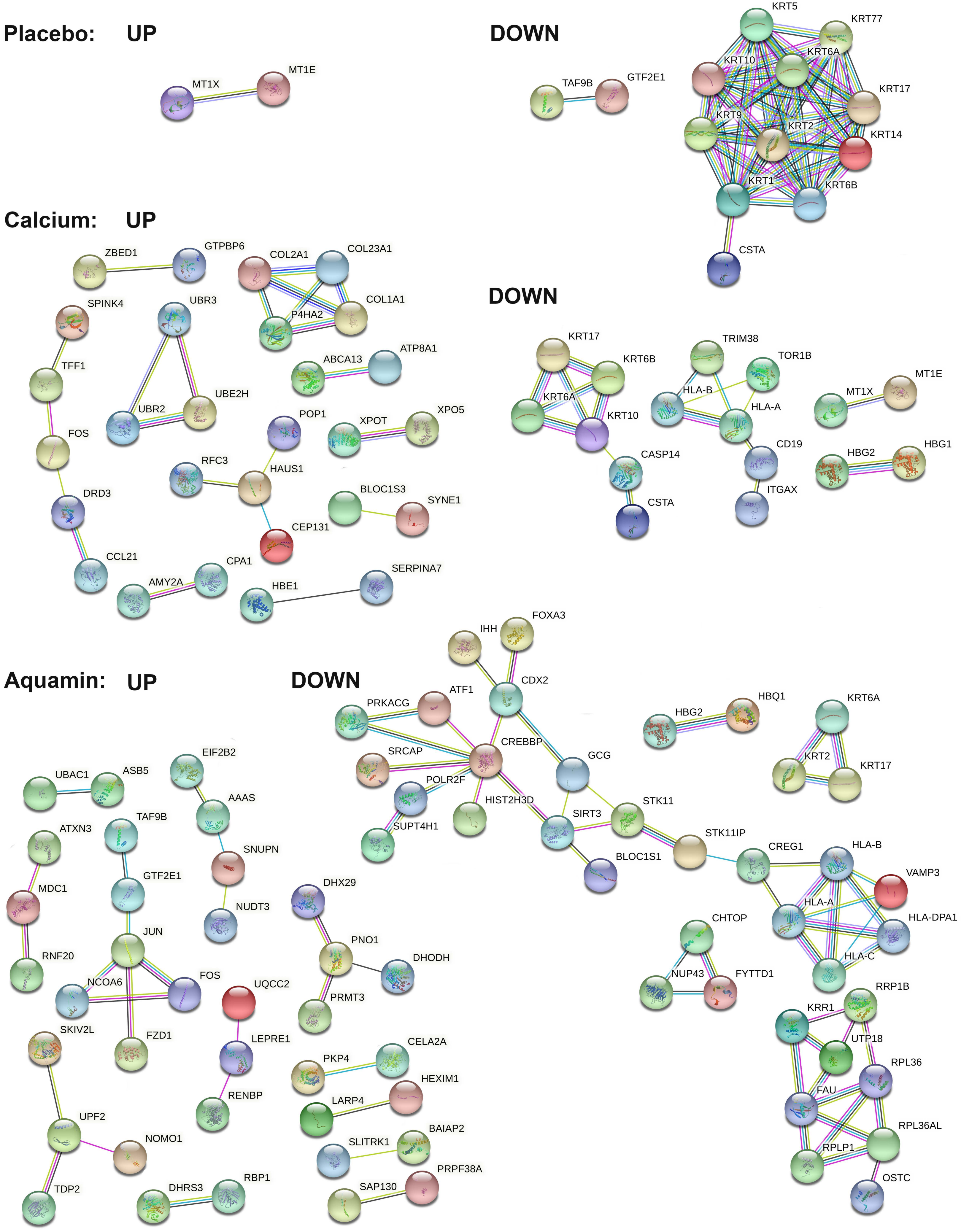

### Supplement Figure 3

#### Significantly Altered Pathways

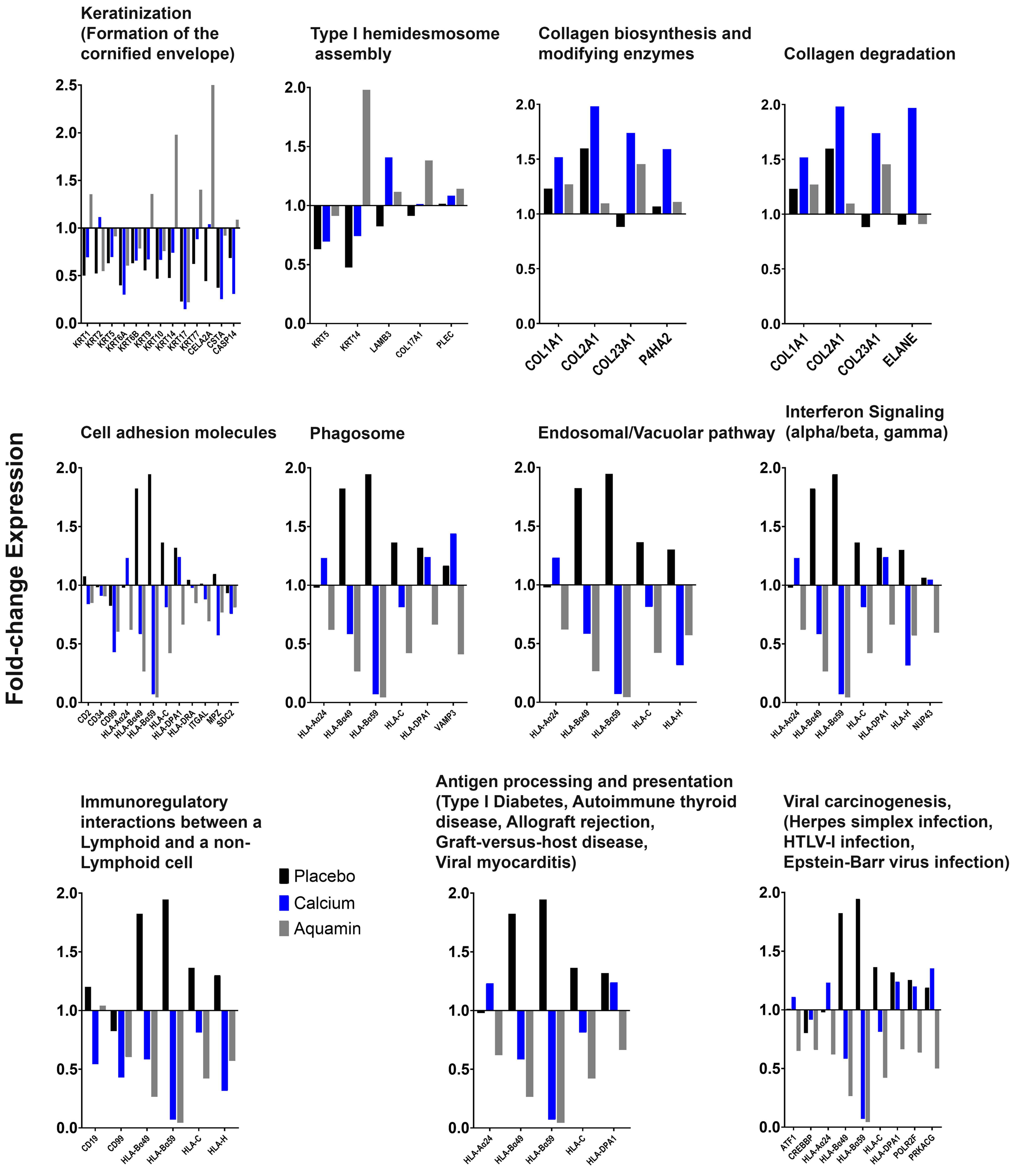
