## Supplement Table 1 for "A MULTI-MINERAL INTERVENTION TO MODULATE COLONIC MUCOSAL PROTEIN PROFILE: Results from a 90-day trial in healthy human subjects"

**Supplement Table 1. List of antibodies used for quantitative immunohistochemistry (qIHC)**

| Antibody | Vendor | Catalog # | Dilution | Incubation Time | Retrieval Method |
|---|---|---|---|---|---|
| Rb Ki-67 mAb clone SP6 | Cell Marque | 275R-16 | 1:250 | 30 min | [a]HIER pH 9.0, 20 min |
| Ms CK20 mAb clone $K_s$20.8 | Dako | M7019 | 1:100 | 60 min | FLEX TRS High pH 9.0, 20 min |
| Ms p21 (WAF1) mAb clone 4D10 | Novocastra (Leica) | NCL-L-WAF-1 | 1:40 | 120 min | [a]HIER pH 6.0, 40 min |

[a]HIER: Heat induced epitope retrieval 10 mM Tris HCl/1 mM EDTA buffer

These antibodies are used for immunohistochemical (IHC) staining of formalin-fixed, paraffin-embedded (FFPE) colon tissue sections.

Positive and negative control tissues (or negative control reagent) ran simultaneously using the same protocol as the actual colon specimens.
