## Supplement Table 2 for "A MULTI-MINERAL INTERVENTION TO MODULATE COLONIC MUCOSAL PROTEIN PROFILE: Results from a 90-day trial in healthy human subjects"

**Supplement Table 2. Unbiased approach: Upregulated moieties.**

| Protein (Gene) | Placebo | Calcium | Aquamin | Calcium (Compared to Placebo) | Aquamin (Compared to Placebo) |
|---|---|---|---|---|---|
| *Common in all 3 groups (1)* | | | | | |
| DENND2A | 1.505 | 1.712 | 3.447 | 1.138 | 2.291 |
| *Common between Placebo and Calcium groups (1)* | | | | | |
| COL2A1 | 1.597 | 1.981 | 1.096 | 1.240 | 0.686 |
| *Common between Calcium and Aquamin groups (7)* | | | | | |
| ZBED1 | 0.991 | 1.824 | 1.717 | 1.840 | 1.732 |
| PKP4 | 1.104 | 1.693 | 1.798 | 1.533 | 1.628 |
| FOS | 0.846 | 3.462 | 1.529 | 4.093 | 1.808 |
| MSRB1 | 1.319 | 2.431 | 1.957 | 1.842 | 1.483 |
| TIMM10B | 1.280 | 1.703 | 1.518 | 1.331 | 1.186 |
| SLITRK1 | 1.498 | 1.667 | 1.643 | 1.113 | 1.097 |
| PDSS2 | 1.153 | 1.678 | 1.604 | 1.454 | 1.391 |
| *Unique to Aquamin (90)* | | | | | |
| ZNF839 | 0.683 | 0.933 | 5.934 | 1.365 | 8.685 |
| CHURC1 | 0.456 | 0.494 | 2.788 | 1.084 | 6.118 |
| BFSP2 | 0.252 | 0.370 | 2.765 | 1.468 | 10.984 |
| ESRRA | 0.366 | 0.462 | 2.632 | 1.262 | 7.195 |
| CELA2A | 0.442 | 1.039 | 2.565 | 2.354 | 5.809 |
| HLA-B α37 | 0.202 | 1.323 | 2.364 | 6.560 | 11.721 |
| SERF2 | 0.524 | 0.626 | 2.340 | 1.194 | 4.463 |
| TAF9B | 0.624 | 1.011 | 2.188 | 1.621 | 3.506 |
| PRPF38A | 0.719 | 0.788 | 2.188 | 1.096 | 3.043 |
| NDUFAF3 | 0.952 | 1.212 | 2.182 | 1.273 | 2.293 |
| GLCE | 0.813 | 0.859 | 2.176 | 1.057 | 2.677 |
| PMVK | 0.896 | 0.896 | 2.146 | 1.000 | 2.395 |
| GTF2E1 | 0.626 | 0.716 | 2.130 | 1.143 | 3.403 |
| ATXN3 | 0.491 | 0.465 | 2.104 | 0.949 | 4.288 |
| NOMO1 | 0.873 | 1.366 | 2.019 | 1.565 | 2.313 |
| TPK1 | 0.723 | 0.677 | 2.012 | 0.936 | 2.784 |
| RNF20 | 0.732 | 0.790 | 2.009 | 1.079 | 2.744 |
| KRT14 | 0.475 | 0.741 | 1.980 | 1.561 | 4.170 |
| FAAH | 0.789 | 0.742 | 1.977 | 0.940 | 2.505 |
| ASB5 | 0.839 | 0.933 | 1.969 | 1.113 | 2.347 |
| FZD1 | 0.879 | 0.908 | 1.930 | 1.033 | 2.196 |
| PITPNM1 | 0.964 | 0.854 | 1.906 | 0.886 | 1.977 |
| DAAM2 | 0.718 | 0.906 | 1.897 | 1.262 | 2.644 |
| SAMD13 | 0.883 | 0.976 | 1.896 | 1.105 | 2.148 |
| FAM174A | 0.549 | 0.532 | 1.865 | 0.968 | 3.393 |
| OTULIN | 0.752 | 0.769 | 1.835 | 1.023 | 2.439 |
| BAIAP2 | 0.527 | 0.840 | 1.810 | 1.594 | 3.435 |
| NECTIN1 | 0.896 | 0.702 | 1.782 | 0.783 | 1.989 |
| SRGAP2C | 0.785 | 0.767 | 1.779 | 0.976 | 2.265 |
| PNO1 | 0.731 | 0.870 | 1.774 | 1.190 | 2.427 |
| ZNF703 | 0.746 | 1.068 | 1.774 | 1.431 | 2.377 |
| TMBIM1 | 0.781 | 0.825 | 1.765 | 1.057 | 2.259 |
| LARP4 | 1.039 | 0.918 | 1.756 | 0.884 | 1.690 |
| CAPN14 | 0.481 | 0.742 | 1.751 | 1.543 | 3.640 |

| | | | | | |
|---|---|---|---|---|---|
| SLC12A7 | 0.644 | 0.781 | 1.749 | 1.212 | 2.713 |
| SKIV2L | 0.833 | 0.711 | 1.747 | 0.855 | 2.098 |
| QTRT2 | 0.836 | 0.973 | 1.735 | 1.164 | 2.076 |
| BCHE | 0.852 | 0.805 | 1.731 | 0.946 | 2.032 |
| FAM109B | 0.497 | 1.112 | 1.719 | 2.238 | 3.459 |
| TECPR1 | 0.908 | 1.008 | 1.710 | 1.110 | 1.883 |
| PACSIN3 | 0.662 | 0.799 | 1.710 | 1.207 | 2.583 |
| MDC1 | 0.875 | 1.210 | 1.692 | 1.383 | 1.934 |
| SLC37A3 | 1.088 | 1.170 | 1.690 | 1.076 | 1.553 |
| UQCC2 | 0.797 | 0.839 | 1.684 | 1.053 | 2.113 |
| SLC41A3 | 1.022 | 0.849 | 1.683 | 0.831 | 1.648 |
| AAAS | 0.831 | 1.000 | 1.682 | 1.203 | 2.022 |
| JUN | 0.801 | 0.672 | 1.680 | 0.839 | 2.098 |
| TDP2 | 0.793 | 0.867 | 1.678 | 1.094 | 2.116 |
| HEXIM1 | 0.792 | 0.979 | 1.676 | 1.236 | 2.116 |
| SLC39A11 | 0.536 | 1.007 | 1.675 | 1.878 | 3.122 |
| ATL2 | 0.707 | 0.780 | 1.674 | 1.103 | 2.369 |
| SKAP2 | 0.882 | 1.007 | 1.660 | 1.142 | 1.882 |
| FRG1 | 0.592 | 0.733 | 1.658 | 1.238 | 2.801 |
| DHODH | 0.872 | 0.854 | 1.654 | 0.980 | 1.898 |
| UPF2 | 0.816 | 0.930 | 1.649 | 1.139 | 2.020 |
| BIRC6 | 0.899 | 0.958 | 1.647 | 1.066 | 1.831 |
| EIF2B2 | 0.994 | 0.692 | 1.629 | 0.696 | 1.639 |
| DHX29 | 0.877 | 0.981 | 1.628 | 1.119 | 1.857 |
| FER1L6 | 0.926 | 1.023 | 1.610 | 1.104 | 1.738 |
| ALDH1L1 | 0.801 | 0.813 | 1.609 | 1.015 | 2.008 |
| TMEM106B | 0.891 | 1.064 | 1.609 | 1.194 | 1.805 |
| DHRS3 | 0.845 | 0.785 | 1.601 | 0.928 | 1.894 |
| RAB3B | 0.857 | 0.857 | 1.590 | 1.000 | 1.856 |
| SOX11 | 1.062 | 1.046 | 1.589 | 0.985 | 1.496 |
| P3H1 | 0.730 | 0.829 | 1.588 | 1.136 | 2.175 |
| COX16 | 0.809 | 0.885 | 1.583 | 1.093 | 1.955 |
| IGKV1-12 | 0.984 | 0.850 | 1.581 | 0.864 | 1.607 |
| WASH6P | 0.728 | 1.069 | 1.577 | 1.469 | 2.166 |
| CHCHD6 | 0.830 | 1.061 | 1.575 | 1.278 | 1.898 |
| TMSB15B | 0.854 | 0.804 | 1.570 | 0.941 | 1.838 |
| PRMT3 | 1.337 | 1.134 | 1.564 | 0.848 | 1.170 |
| SULT1C2 | 0.906 | 0.961 | 1.562 | 1.060 | 1.724 |
| CEP162 | 1.004 | 0.880 | 1.558 | 0.877 | 1.551 |
| HEATR3 | 0.860 | 0.921 | 1.551 | 1.071 | 1.804 |
| RENBP | 1.034 | 1.000 | 1.544 | 0.966 | 1.492 |
| CMTM5 | 0.939 | 0.773 | 1.534 | 0.823 | 1.634 |
| FITM2 | 0.766 | 0.851 | 1.531 | 1.111 | 1.998 |
| MED20 | 0.958 | 0.801 | 1.530 | 0.835 | 1.596 |
| CARD16 | 0.741 | 0.797 | 1.529 | 1.074 | 2.062 |
| TASP1 | 0.754 | 0.859 | 1.528 | 1.139 | 2.026 |
| SLC35A2 | 0.541 | 0.697 | 1.527 | 1.288 | 2.822 |
| NCOA6 | 0.673 | 0.895 | 1.527 | 1.329 | 2.267 |
| SNUPN | 1.058 | 1.083 | 1.526 | 1.024 | 1.443 |
| KAT8 | 0.840 | 0.867 | 1.522 | 1.031 | 1.811 |
| ARID2 | 0.784 | 0.784 | 1.521 | 1.000 | 1.940 |

| | | | | | |
|---|---|---|---|---|---|
| NUDT3 | 0.813 | 0.765 | 1.514 | 0.942 | 1.863 |
| UBAC1 | 0.829 | 0.922 | 1.505 | 1.111 | 1.815 |
| RBP1 | 0.931 | 0.848 | 1.503 | 0.910 | 1.614 |
| TMEM222 | 0.925 | 0.803 | 1.498 | 0.868 | 1.618 |
| SAP130 | 0.828 | 0.816 | 1.495 | 0.985 | 1.805 |
| | | *Unique to Calcium (66)* | | | |
| DRD3 | 0.640 | 6.746 | 1.129 | 10.544 | 1.764 |
| UBR3 | 0.871 | 3.384 | 1.126 | 3.884 | 1.293 |
| CPA1 | 1.217 | 3.238 | 1.030 | 2.659 | 0.846 |
| ADAD1 | 0.706 | 2.806 | 1.365 | 3.974 | 1.933 |
| GPATCH1 | 0.566 | 2.774 | 1.029 | 4.899 | 1.817 |
| RPUSD4 | 0.774 | 2.647 | 0.876 | 3.419 | 1.132 |
| BLOC1S3 | 1.174 | 2.595 | 1.247 | 2.211 | 1.062 |
| SLC22A11 | 1.289 | 2.473 | 1.247 | 1.919 | 0.968 |
| CD101 | 0.916 | 2.451 | 1.302 | 2.675 | 1.421 |
| TXNDC15 | 1.035 | 2.365 | 1.004 | 2.284 | 0.970 |
| HBE1 | 0.791 | 2.302 | 0.954 | 2.910 | 1.206 |
| FAT3 | 1.007 | 2.276 | 1.147 | 2.261 | 1.139 |
| BCR | 0.892 | 2.185 | 1.195 | 2.451 | 1.340 |
| CCL21 | 0.940 | 2.163 | 1.125 | 2.300 | 1.197 |
| RARRES1 | 0.903 | 2.108 | 1.341 | 2.335 | 1.485 |
| O43824 | 0.927 | 2.048 | 1.335 | 2.209 | 1.440 |
| LENG1 | 0.905 | 2.044 | 1.218 | 2.259 | 1.346 |
| POP1 | 0.925 | 1.999 | 1.235 | 2.161 | 1.335 |
| HAUS1 | 0.971 | 1.993 | 1.311 | 2.054 | 1.351 |
| ATRN | 0.944 | 1.971 | 1.248 | 2.087 | 1.321 |
| CEP131 | 0.943 | 1.970 | 1.105 | 2.088 | 1.172 |
| ELANE | 0.903 | 1.968 | 0.909 | 2.179 | 1.007 |
| DHX30 | 0.755 | 1.901 | 1.125 | 2.520 | 1.490 |
| AMY2A | 1.172 | 1.872 | 0.517 | 1.598 | 0.441 |
| CROT | 1.004 | 1.868 | 0.831 | 1.861 | 0.827 |
| PTBP3 | 0.936 | 1.846 | 0.790 | 1.972 | 0.844 |
| NCKAP5L | 0.878 | 1.811 | 1.153 | 2.063 | 1.313 |
| CAPS | 0.919 | 1.796 | 1.065 | 1.955 | 1.159 |
| DNAJC1 | 0.846 | 1.784 | 0.802 | 2.109 | 0.948 |
| RNMT | 0.996 | 1.778 | 1.127 | 1.785 | 1.131 |
| ECHDC3 | 0.873 | 1.777 | 1.046 | 2.035 | 1.198 |
| HLA-A α31 | 0.797 | 1.773 | 0.856 | 2.224 | 1.074 |
| ABCA13 | 0.988 | 1.747 | 0.975 | 1.768 | 0.988 |
| COL23A1 | 0.882 | 1.738 | 1.454 | 1.971 | 1.649 |
| SMG5 | 0.919 | 1.732 | 1.024 | 1.885 | 1.115 |
| CPSF2 | 1.032 | 1.724 | 1.071 | 1.671 | 1.038 |
| SPINK4 | 0.997 | 1.722 | 0.901 | 1.727 | 0.904 |
| SLC35B3 | 1.034 | 1.721 | 1.135 | 1.664 | 1.097 |
| FAT1 | 0.937 | 1.717 | 1.454 | 1.833 | 1.552 |
| SYNE1 | 0.996 | 1.686 | 1.363 | 1.694 | 1.369 |
| DUSP16 | 0.862 | 1.683 | 1.039 | 1.953 | 1.205 |
| HSP90AA4P | 0.969 | 1.679 | 1.093 | 1.733 | 1.128 |
| AKAP5 | 1.021 | 1.661 | 1.083 | 1.627 | 1.061 |
| PKLR | 0.769 | 1.654 | 1.267 | 2.151 | 1.648 |
| TRPM3 | 0.999 | 1.636 | 1.438 | 1.638 | 1.439 |

| | | | | | |
|---|---|---|---|---|---|
| XPO5 | 1.033 | 1.634 | 1.027 | 1.582 | 0.995 |
| METTL13 | 1.191 | 1.605 | 1.179 | 1.347 | 0.990 |
| WASHC3 | 1.171 | 1.604 | 0.885 | 1.370 | 0.755 |
| PCIF1 | 0.892 | 1.604 | 1.039 | 1.798 | 1.165 |
| UBR2 | 1.011 | 1.602 | 1.062 | 1.585 | 1.050 |
| MRPL54 | 1.062 | 1.595 | 1.020 | 1.501 | 0.960 |
| P4HA2 | 1.068 | 1.590 | 1.109 | 1.488 | 1.038 |
| ATP8A1 | 1.026 | 1.579 | 0.964 | 1.539 | 0.940 |
| TSPAN1 | 1.047 | 1.565 | 1.033 | 1.494 | 0.987 |
| WDR98 | 1.235 | 1.556 | 1.166 | 1.260 | 0.944 |
| GGT2 | 1.018 | 1.554 | 1.313 | 1.527 | 1.290 |
| UBE2H | 1.228 | 1.545 | 0.835 | 1.258 | 0.680 |
| XPOT | 0.977 | 1.543 | 1.380 | 1.579 | 1.413 |
| SERPINA7 | 1.063 | 1.542 | 0.750 | 1.450 | 0.705 |
| TFF1 | 1.215 | 1.536 | 1.062 | 1.265 | 0.874 |
| EMC6 | 1.082 | 1.522 | 1.013 | 1.406 | 0.936 |
| COL1A1 | 1.230 | 1.516 | 1.271 | 1.232 | 1.033 |
| STAT4 | 0.970 | 1.512 | 1.233 | 1.559 | 1.272 |
| CNOT8 | 0.956 | 1.511 | 1.161 | 1.580 | 1.213 |
| STAB1 | 1.003 | 1.504 | 1.107 | 1.500 | 1.104 |
| RFC3 | 0.847 | 1.503 | 1.136 | 1.775 | 1.341 |
| *Unique to Placebo (10)* | | | | | |
| HLA-B α59 | 1.946 | 0.070 | 0.044 | 0.036 | 0.023 |
| HLA-B α49 | 1.823 | 0.584 | 0.265 | 0.320 | 0.145 |
| MT1E | 1.747 | 0.540 | 1.492 | 0.309 | 0.854 |
| SUPT4H1 | 1.665 | 1.421 | 0.233 | 0.853 | 0.140 |
| CMTM6 | 1.583 | 1.136 | 0.183 | 0.717 | 0.116 |
| MT1X | 1.578 | 0.580 | 0.675 | 0.368 | 0.427 |
| CTNNBIP1 | 1.521 | 1.058 | 0.748 | 0.696 | 0.492 |
| FAM84B | 1.520 | 1.237 | 0.339 | 0.814 | 0.223 |
| HECW2 | 1.516 | 0.957 | 0.552 | 0.631 | 0.365 |
| NAT1 | 1.503 | 0.922 | 0.587 | 0.614 | 0.390 |

The values represent fold-change of abundance ratio as compared to the control (all baseline samples and post-intervention placebo samples). These samples were assessed by TMT based differential proteomic expression by pooling the samples for each group. Protein FDR Confidence for all the proteins was <2%. Proteins were upregulated by 1.5-fold as compared to control. These data are also presented in Figure 3A and 3C.
