## Supplement Table 3 for "A MULTI-MINERAL INTERVENTION TO MODULATE COLONIC MUCOSAL PROTEIN PROFILE: Results from a 90-day trial in healthy human subjects"

**Supplement Table 3. Unbiased approach: Downregulated moieties.**

| Protein (Gene) | Placebo | Calcium | Aquamin | Calcium (Compared to Placebo) | Aquamin (Compared to Placebo) |
|---|---|---|---|---|---|
| *Common in all 3 groups (3)* | | | | | |
| KRT6A | 0.398 | 0.302 | 0.606 | 0.757 | 1.520 |
| KRT17 | 0.228 | 0.149 | 0.220 | 0.653 | 0.967 |
| MTMR11 | 0.665 | 0.576 | 0.557 | 0.867 | 0.837 |
| *Common between Placebo and Calcium groups (10)* | | | | | |
| BFSP2 | 0.252 | 0.370 | 2.765 | 1.468 | 10.984 |
| CHURC1 | 0.456 | 0.494 | 2.788 | 1.084 | 6.118 |
| ESRRA | 0.366 | 0.462 | 2.632 | 1.262 | 7.195 |
| SERPINB4 | 0.435 | 0.225 | 1.086 | 0.518 | 2.498 |
| KRT6B | 0.630 | 0.657 | 0.785 | 1.042 | 1.246 |
| SERF2 | 0.524 | 0.626 | 2.340 | 1.194 | 4.463 |
| CSTA | 0.374 | 0.254 | 0.920 | 0.679 | 2.457 |
| FAM174A | 0.549 | 0.532 | 1.865 | 0.968 | 3.393 |
| ATXN3 | 0.491 | 0.465 | 2.104 | 0.949 | 4.288 |
| KRT10 | 0.467 | 0.665 | 0.758 | 1.424 | 1.622 |
| *Common between Calcium and Aquamin groups (6)* | | | | | |
| CD99 | 0.824 | 0.429 | 0.604 | 0.521 | 0.733 |
| HLA-B α49 | 1.823 | 0.584 | 0.265 | 0.320 | 0.145 |
| HACD2 | 0.853 | 0.669 | 0.548 | 0.785 | 0.642 |
| HLA-H | 1.300 | 0.315 | 0.571 | 0.242 | 0.439 |
| HBG2 | 1.021 | 0.473 | 0.372 | 0.463 | 0.365 |
| HLA-B α59 | 1.946 | 0.070 | 0.044 | 0.036 | 0.023 |
| *Common between Placebo and Aquamin groups (4)* | | | | | |
| FBN1 | 0.515 | 0.734 | 0.655 | 1.426 | 1.272 |
| ZNF280D | 0.557 | 1.182 | 0.611 | 2.122 | 1.098 |
| KRT2 | 0.524 | 1.116 | 0.549 | 2.131 | 1.048 |
| CEP170B | 0.618 | 0.844 | 0.612 | 1.366 | 0.990 |
| *Unique to Aquamin (78)* | | | | | |
| ZNF33B | 1.177 | 1.344 | 0.172 | 1.142 | 0.146 |
| CMTM6 | 1.583 | 1.136 | 0.183 | 0.717 | 0.116 |
| UTP18 | 1.248 | 1.323 | 0.184 | 1.060 | 0.148 |
| SUPT4H1 | 1.665 | 1.421 | 0.233 | 0.853 | 0.140 |
| SPATA2 | 1.402 | 1.306 | 0.296 | 0.931 | 0.211 |
| CREG1 | 1.470 | 1.416 | 0.320 | 0.963 | 0.218 |
| FAM84B | 1.520 | 1.237 | 0.339 | 0.814 | 0.223 |
| MLXIP | 1.120 | 1.032 | 0.377 | 0.921 | 0.337 |
| VAMP3 | 1.167 | 1.440 | 0.412 | 1.233 | 0.353 |
| CRYZL1 | 1.119 | 1.244 | 0.412 | 1.112 | 0.368 |
| HLA-C | 1.364 | 0.813 | 0.421 | 0.596 | 0.309 |
| RPL36 | 1.254 | 0.954 | 0.423 | 0.761 | 0.338 |
| FOXA3 | 0.980 | 0.852 | 0.430 | 0.869 | 0.439 |
| SPG21 | 0.998 | 0.705 | 0.459 | 0.706 | 0.460 |
| FAU | 1.347 | 1.038 | 0.482 | 0.771 | 0.358 |
| PNLIPRP2 | 1.016 | 0.944 | 0.483 | 0.929 | 0.476 |
| SNX17 | 1.195 | 1.134 | 0.492 | 0.949 | 0.412 |
| PRKACG | 1.189 | 1.354 | 0.500 | 1.139 | 0.421 |
| CCL15 | 1.479 | 1.323 | 0.510 | 0.895 | 0.345 |

| | | | | | |
|---|---|---|---|---|---|
| AMY2A | 1.172 | 1.872 | 0.517 | 1.598 | 0.441 |
| BLOC1S1 | 1.232 | 1.237 | 0.519 | 1.004 | 0.421 |
| AMIGO1 | 1.162 | 1.028 | 0.521 | 0.885 | 0.449 |
| TPCN1 | 1.104 | 0.953 | 0.531 | 0.863 | 0.481 |
| HBQ1 | 0.878 | 1.176 | 0.539 | 1.340 | 0.614 |
| VMP1 | 1.270 | 0.985 | 0.539 | 0.776 | 0.425 |
| AASS | 1.488 | 1.128 | 0.540 | 0.758 | 0.363 |
| DAZAP1 | 0.992 | 0.981 | 0.542 | 0.989 | 0.547 |
| HIST2H3A | 1.057 | 1.021 | 0.544 | 0.966 | 0.514 |
| EMC3 | 1.200 | 1.164 | 0.546 | 0.970 | 0.455 |
| HECW2 | 1.516 | 0.957 | 0.552 | 0.631 | 0.365 |
| ADARB1 | 1.073 | 0.912 | 0.554 | 0.850 | 0.516 |
| RNF2 | 1.133 | 1.021 | 0.557 | 0.901 | 0.491 |
| OSTC | 1.175 | 0.845 | 0.558 | 0.719 | 0.475 |
| TMEM263 | 1.064 | 1.312 | 0.567 | 1.233 | 0.532 |
| STK11 | 0.910 | 0.858 | 0.581 | 0.943 | 0.639 |
| RNASE6 | 1.227 | 1.054 | 0.585 | 0.860 | 0.477 |
| NAT1 | 1.503 | 0.922 | 0.587 | 0.614 | 0.390 |
| CHTOP | 1.049 | 1.086 | 0.587 | 1.035 | 0.560 |
| GCG | 0.964 | 0.906 | 0.591 | 0.939 | 0.613 |
| ATP5J2 | 1.180 | 0.990 | 0.595 | 0.838 | 0.504 |
| NUP43 | 1.063 | 1.048 | 0.597 | 0.986 | 0.562 |
| ARHGAP21 | 1.263 | 1.079 | 0.598 | 0.854 | 0.474 |
| RBBP9 | 1.224 | 1.136 | 0.599 | 0.928 | 0.490 |
| ZMYM4 | 1.156 | 1.094 | 0.603 | 0.947 | 0.522 |
| SMC1B | 1.146 | 1.140 | 0.610 | 0.994 | 0.532 |
| PCM1 | 0.788 | 0.988 | 0.613 | 1.254 | 0.778 |
| HLA-A | 0.979 | 1.233 | 0.620 | 1.259 | 0.633 |
| IGLV2-14 | 1.250 | 1.262 | 0.623 | 1.010 | 0.498 |
| LIMD2 | 1.122 | 0.830 | 0.624 | 0.740 | 0.556 |
| KRR1 | 1.212 | 1.079 | 0.627 | 0.891 | 0.518 |
| DHRS13 | 0.865 | 0.876 | 0.628 | 1.013 | 0.726 |
| NOX1 | 1.186 | 1.228 | 0.632 | 1.036 | 0.533 |
| POLR2F | 1.254 | 1.200 | 0.637 | 0.957 | 0.508 |
| CDX1 | 0.702 | 0.671 | 0.638 | 0.955 | 0.908 |
| CAPN13 | 0.825 | 0.748 | 0.638 | 0.907 | 0.773 |
| TANK | 1.012 | 1.059 | 0.639 | 1.046 | 0.631 |
| STK11IP | 1.240 | 1.229 | 0.648 | 0.991 | 0.522 |
| SRCAP | 1.001 | 0.842 | 0.648 | 0.842 | 0.647 |
| MISP3 | 1.143 | 0.930 | 0.648 | 0.813 | 0.567 |
| FYTTD1 | 1.033 | 1.007 | 0.648 | 0.974 | 0.627 |
| DDX3Y | 0.965 | 0.740 | 0.649 | 0.767 | 0.672 |
| ATF1 | 0.997 | 1.110 | 0.650 | 1.113 | 0.652 |
| LMBRD2 | 1.059 | 1.149 | 0.651 | 1.085 | 0.615 |
| ACOT1 | 1.203 | 0.994 | 0.654 | 0.826 | 0.544 |
| CREBBP | 0.802 | 0.916 | 0.658 | 1.142 | 0.820 |
| ISCA1 | 1.204 | 1.117 | 0.658 | 0.928 | 0.546 |
| RRP1B | 0.881 | 1.281 | 0.660 | 1.454 | 0.749 |
| PLEKHA1 | 1.224 | 1.093 | 0.661 | 0.893 | 0.541 |
| TOMM5 | 1.076 | 1.048 | 0.662 | 0.973 | 0.615 |
| IHH | 1.210 | 1.166 | 0.662 | 0.963 | 0.547 |

| | | | | | |
|---|---|---|---|---|---|
| CDX2 | 0.903 | 1.072 | 0.664 | 1.187 | 0.736 |
| HLA-DPA1 | 1.319 | 1.241 | 0.664 | 0.941 | 0.504 |
| SIRT3 | 1.176 | 1.222 | 0.664 | 1.040 | 0.565 |
| RPL36AL | 1.234 | 0.952 | 0.665 | 0.771 | 0.539 |
| ELMSAN1 | 0.986 | 1.082 | 0.666 | 1.097 | 0.676 |
| AKR1C1 | 1.131 | 1.034 | 0.668 | 0.914 | 0.590 |
| CCNYL1 | 1.074 | 1.065 | 0.668 | 0.992 | 0.622 |
| RPLP1 | 1.177 | 1.058 | 0.668 | 0.899 | 0.568 |
| | | *Unique to Calcium (45)* | | | |
| CASP14 | 0.684 | 0.308 | 1.087 | 0.450 | 1.590 |
| ARF3 | 1.103 | 0.319 | 0.839 | 0.289 | 0.760 |
| UAP1L1 | 0.888 | 0.367 | 0.934 | 0.414 | 1.052 |
| CTNNA2 | 0.994 | 0.431 | 0.957 | 0.434 | 0.962 |
| HSPA6 | 0.754 | 0.479 | 1.493 | 0.636 | 1.979 |
| VWA5B1 | 1.139 | 0.483 | 1.097 | 0.424 | 0.963 |
| ERCC6 | 0.767 | 0.516 | 1.126 | 0.672 | 1.467 |
| PARVG | 1.062 | 0.517 | 0.909 | 0.487 | 0.856 |
| TRIM38 | 0.850 | 0.520 | 0.786 | 0.611 | 0.924 |
| NCALD | 1.018 | 0.528 | 0.909 | 0.519 | 0.894 |
| MT1E | 1.747 | 0.540 | 1.492 | 0.309 | 0.854 |
| CD19 | 1.202 | 0.544 | 1.041 | 0.453 | 0.866 |
| DCP1B | 0.828 | 0.545 | 0.905 | 0.658 | 1.092 |
| NAA20 | 0.934 | 0.547 | 0.877 | 0.586 | 0.939 |
| NACA2 | 0.937 | 0.556 | 0.973 | 0.593 | 1.039 |
| GON7 | 0.839 | 0.568 | 0.776 | 0.677 | 0.925 |
| MPZ | 1.096 | 0.573 | 0.768 | 0.523 | 0.701 |
| RANBP6 | 0.959 | 0.575 | 0.973 | 0.600 | 1.014 |
| SPATA5L1 | 0.795 | 0.579 | 1.020 | 0.728 | 1.283 |
| MT1X | 1.578 | 0.580 | 0.675 | 0.368 | 0.427 |
| TOGARAM2 | 0.996 | 0.583 | 0.876 | 0.585 | 0.879 |
| TSG101 | 0.955 | 0.583 | 0.967 | 0.611 | 1.013 |
| RPS4Y2 | 1.224 | 0.586 | 0.767 | 0.478 | 0.626 |
| TOR1B | 1.119 | 0.598 | 0.919 | 0.534 | 0.821 |
| HBG1 | 0.882 | 0.600 | 0.842 | 0.681 | 0.955 |
| PTBP2 | 0.748 | 0.601 | 1.148 | 0.803 | 1.534 |
| AOAH | 1.171 | 0.607 | 0.897 | 0.519 | 0.766 |
| RNF181 | 1.097 | 0.610 | 0.747 | 0.556 | 0.681 |
| INSL5 | 0.807 | 0.614 | 0.994 | 0.760 | 1.232 |
| VPS72 | 0.926 | 0.617 | 0.901 | 0.667 | 0.973 |
| DOPEY1 | 1.150 | 0.621 | 1.178 | 0.540 | 1.024 |
| HLA-A | 1.385 | 0.633 | 0.795 | 0.457 | 0.574 |
| PIGU | 0.972 | 0.633 | 0.939 | 0.651 | 0.966 |
| PPWD1 | 0.970 | 0.634 | 0.843 | 0.654 | 0.868 |
| C2orf72 | 1.055 | 0.641 | 0.925 | 0.607 | 0.876 |
| ITGAX | 0.935 | 0.641 | 1.367 | 0.685 | 1.461 |
| OSBPL3 | 0.946 | 0.641 | 0.989 | 0.678 | 1.046 |
| SEPHS2 | 0.903 | 0.645 | 1.206 | 0.714 | 1.335 |
| PIGBOS1 | 0.959 | 0.649 | 1.233 | 0.676 | 1.285 |
| F8A1 | 1.020 | 0.649 | 0.908 | 0.637 | 0.890 |
| COX11 | 1.029 | 0.657 | 1.247 | 0.638 | 1.213 |
| FADD | 0.857 | 0.663 | 1.072 | 0.774 | 1.251 |

| | | | | | |
|---|---|---|---|---|---|
| ACSM4 | 1.083 | 0.663 | 0.774 | 0.612 | 0.714 |
| PHF23 | 0.709 | 0.668 | 0.729 | 0.942 | 1.029 |
| TRABD | 0.964 | 0.668 | 0.980 | 0.693 | 1.016 |
| | | *Unique to Placebo (31)* | | | |
| HLA-B α37 | 0.202 | 1.323 | 2.364 | 6.560 | 11.721 |
| CELA2A | 0.442 | 1.039 | 2.565 | 2.354 | 5.809 |
| HSF1 | 0.467 | 0.884 | 0.842 | 1.891 | 1.802 |
| KRT14 | 0.475 | 0.741 | 1.980 | 1.561 | 4.170 |
| CAPN14 | 0.481 | 0.742 | 1.751 | 1.543 | 3.640 |
| DCD | 0.481 | 0.716 | 0.815 | 1.488 | 1.695 |
| FAM109B | 0.497 | 1.112 | 1.719 | 2.238 | 3.459 |
| KRT1 | 0.500 | 0.694 | 1.355 | 1.387 | 2.710 |
| BAIAP2 | 0.527 | 0.840 | 1.810 | 1.594 | 3.435 |
| SLC39A11 | 0.536 | 1.007 | 1.675 | 1.878 | 3.122 |
| SLC35A2 | 0.541 | 0.697 | 1.527 | 1.288 | 2.822 |
| MYH10 | 0.543 | 0.988 | 1.388 | 1.821 | 2.558 |
| KRT9 | 0.555 | 0.671 | 1.357 | 1.209 | 2.445 |
| GPATCH1 | 0.566 | 2.774 | 1.029 | 4.899 | 1.817 |
| NKRF | 0.567 | 1.093 | 1.110 | 1.930 | 1.960 |
| FRG1 | 0.592 | 0.733 | 1.658 | 1.238 | 2.801 |
| CELA3A | 0.610 | 1.017 | 0.746 | 1.667 | 1.222 |
| ZSWIM8 | 0.614 | 1.289 | 1.252 | 2.099 | 2.040 |
| KRT77 | 0.622 | 0.882 | 1.401 | 1.419 | 2.254 |
| TAF9B | 0.624 | 1.011 | 2.188 | 1.621 | 3.506 |
| GTF2E1 | 0.626 | 0.716 | 2.130 | 1.143 | 3.403 |
| KRT5 | 0.631 | 0.695 | 0.912 | 1.102 | 1.446 |
| DRD3 | 0.640 | 6.746 | 1.129 | 10.544 | 1.764 |
| URI1 | 0.641 | 1.458 | 1.395 | 2.273 | 2.176 |
| SLC12A7 | 0.644 | 0.781 | 1.749 | 1.212 | 2.713 |
| C19orf53 | 0.645 | 0.753 | 0.989 | 1.166 | 1.533 |
| UBN2 | 0.655 | 0.819 | 0.976 | 1.250 | 1.491 |
| JPT2 | 0.660 | 0.893 | 1.170 | 1.354 | 1.773 |
| PACSIN3 | 0.662 | 0.799 | 1.710 | 1.207 | 2.583 |
| FAM83F | 0.665 | 0.892 | 1.093 | 1.342 | 1.645 |
| FAM117B | 0.670 | 0.788 | 0.900 | 1.176 | 1.343 |

The values represent fold-change of abundance ratio as compared to the control (all baseline samples and post-intervention placebo samples). These samples were assessed by TMT based differential proteomic expression by pooling the samples for each group. Protein FDR Confidence for all the proteins was <2%. Proteins were downregulated by 1.5-fold as compared to control. These data are also presented in Figure 3A and 3D.
