## Supplemental File 1 for "A MULTI-MINERAL INTERVENTION TO MODULATE COLONIC MUCOSAL PROTEIN PROFILE: Results from a 90-day trial in healthy human subjects"

**Supplemental File 1. Downregulated Proteins of Interest.** (Proteins presented in Figure 5)

Ki67: Ki67 antigen expresses throughout the cell cycle in proliferating cells. A high Ki67 expression is correlated with poor survival, indicating that increased Ki67 expression may serve as a predictive method for poor prognosis of colorectal cancer patients.


- Huh JW, Lee JH, Kim HR. Expression of p16, p53, and Ki-67 in colorectal adenocarcinoma: a study of 356 surgically resected cases. Hepatogastroenterology. 2010;57(101):734-740.
- Luo ZW, Zhu MG, Zhang ZQ, Ye FJ, Huang WH, Luo XZ. Increased expression of Ki-67 is a poor prognostic marker for colorectal cancer patients: a meta-analysis. BMC Cancer. 2019;19(1):123. doi:10.1186/s12885-019-5324-y


CLDN7: Claudin-7 is one of the pore forming claudins and plays a major role in tight junction-specific obliteration of the intercellular space.


- Overgaard CE, Daugherty BL, Mitchell LA, Koval M. Claudins: control of barrier function and regulation in response to oxidant stress. Antioxid Redox Signal. 2011;15(5):1179-1193. doi:10.1089/ars.2011.3893
- Markov AG, Aschenbach JR, Amasheh S. Claudin clusters as determinants of epithelial barrier function. IUBMB Life. 2015;67(1):29-35. doi:10.1002/iub.1347


STAT1: A high protein expression of Signal transducer and activator of transcription 1-alpha/beta (STAT1) in early-stage colorectal cancer, particularly of the MSI subtype, is positively correlated with shorter patient survival times.


- Tanaka A, Zhou Y, Ogawa M, et al. STAT1 as a potential prognosis marker for poor outcomes of early stage colorectal cancer with microsatellite instability. PLoS One. 2020;15(4):e0229252. doi:10.1371/journal.pone.0229252


NOX1: NADPH oxidase 1 is expressed in normal colon epithelial cells and in colon tumor cell lines, and overexpression has been implicated in stimulation of mitogenesis and angiogenesis and inhibition of apoptosis. Additionally, activation of NOX1 through forming a complex with TNF signaling components plays a key role in TNF-induced necrotic cell death.


- Laurent E, McCoy JW 3rd, Macina RA, et al. Nox1 is over-expressed in human colon cancers and correlates with activating mutations in K-Ras. Int J Cancer. 2008;123(1):100-107. doi:10.1002/ijc.23423
- Kim YS, Morgan MJ, Choksi S, Liu ZG. TNF-induced activation of the Nox1 NADPH oxidase and its role in the induction of necrotic cell death. Mol Cell. 2007;26(5):675-687. doi:10.1016/j.molcel.2007.04.021


COX6C: Cytochrome c oxidase subunit 6C (COX6C) is a component of the cytochrome c oxidase, the last enzyme in the mitochondrial electron transport chain which drives oxidative phosphorylation, a pathway that is part of Energy metabolism.


- Yang J, Liu J, Zhang S, Yang Y, Gong J. The overexpression of cytochrome c oxidase subunit 6C activated by Kras mutation is related to energy metabolism in pancreatic cancer. TRANSLATIONAL CANCER RESEARCH. 2018;7(2):290-300.
- Fontanesi F, Soto IC, Horn D, Barrientos A. Assembly of mitochondrial cytochrome c-oxidase, a complicated and highly regulated cellular process. Am J Physiol Cell Physiol. 2006;291(6):C1129-C1147. doi:10.1152/ajpcell.00233.2006
- Herrmann PC, Gillespie JW, Charboneau L, et al. Mitochondrial proteome: altered cytochrome c oxidase subunit levels in prostate cancer. Proteomics. 2003;3(9):1801-1810. doi:10.1002/pmic.200300461


DPAGT1: N-glycosylation of E-cadherin is controlled by the UDP-N-acetylglucosamine-dolichyl-phosphate N-acetylglucosaminephosphotransferase (DPAGT1) gene, a key regulator of the N-glycosylation pathway. DPAGT1 is a target of the canonical Wnt signaling pathway, with both β- and γ-catenins binding to Tcf at its promoter. In a subset of human oral squamous cell carcinoma (OSCC) tumor specimens, overexpression of the DPAGT1 gene, encoding the dolichol-P-dependent N-acetylglucoseamine-1-phosphate transferase, a key regulator of the metabolic pathway of protein N-glycosylation, drives tumor cell discohesion by inhibiting E-cadherin adhesive function.

- Sengupta PK, Bouchie MP, Nita-Lazar M, Yang HY, Kukuruzinska MA. Coordinate regulation of N-glycosylation gene DPAGT1, canonical Wnt signaling and E-cadherin adhesion. J Cell Sci. 2013;126(Pt 2):484-496. doi:10.1242/jcs.113035
- Jamal B, Sengupta PK, Gao ZN, et al. Aberrant amplification of the crosstalk between canonical Wnt signaling and N-glycosylation gene DPAGT1 promotes oral cancer. Oral Oncol. 2012;48(6):523-529. doi:10.1016/j.oraloncology.2012.01.010

VMP1: Vacuole membrane protein 1 is a stress-induced protein that, when overexpressed, promotes formation of intracellular vacuoles followed by cell death, also, known as nanoparticle triggered autophagic cell death. VMP1 is essentially a cell membrane protein and that it is involved in cell–cell adhesion, invasion and metastasis.

- Sauermann M, Sahin O, Sültmann H, et al. Reduced expression of vacuole membrane protein 1 affects the invasion capacity of tumor cells. Oncogene. 2008;27(9):1320-1326. doi:10.1038/sj.onc.1210743
- Amirfallah A, Arason A, Einarsson H, et al. High expression of the vacuole membrane protein 1 (VMP1) is a potential marker of poor prognosis in HER2 positive breast cancer. PLoS One. 2019;14(8):e0221413. doi:10.1371/journal.pone.0221413

CD99: CD99 antigen (CD99) is a cell adhesion molecule and involved in leukocyte transendothelial migration. It is also involved in the lymphocyte and polymorphonuclear leukocytes trafficking and correlated positively with inflammatory bowel disease (IBD) disease activity.

Metastasis requires tumor cells to cross endothelial cell (EC) barriers and this occurs using mechanisms similar to those used by extravasating leukocytes during inflammation. The cell surface receptor CD99 is expressed by leucocytes and EC and participates in inflammatory transendothelial migration (TEM). CD99 is also expressed by tumor cells and have a role in tumor progression and cancer cell TEM.

- Zhou G, Yang W, Yu L, Yu T, Liu Z. CD99 refers to the activity of inflammatory bowel disease. Scand J Gastroenterol. 2017;52(3):359-364. doi:10.1080/00365521.2016.1256426.
- Aarren J. Mannion, Adam F. Odell, Alison Taylor, Pamela F. Jones, Graham P Cook. CD99 regulates cancer cell transendothelial migration and endothelial cell function via CDC42 and actin remodeling. bioRxiv 760934; doi: https://doi.org/10.1101/760934
- Manara MC, Pasello M, Scotlandi K. CD99: A Cell Surface Protein with an Oncojanus Role in Tumors. Genes (Basel). 2018;9(3):159. doi:10.3390/genes9030159

RNF2: The knockdown of E3 ubiquitin-protein ligase RING2 (RNF2) inhibits both cell proliferation and colony formation in soft agar, and induces apoptosis in cancer cells. RNF2 is important for cancer cell survival and proliferation, it could serve as a good target for cancer therapy or prevention. These studies have shown that expression level of p21 elevated by the knockdown of RNF2. In our study, RNF2 is downregulated and CDKN1B (a gene similar to P21) is upregulated with calcium and Aquamin (1.05 with calcium and 1.33 with Aquamin).

- Wen W, Peng C, Kim MO, et al. Knockdown of RNF2 induces apoptosis by regulating MDM2 and p53 stability. Oncogene. 2014;33(4):421-428. doi:10.1038/onc.2012.605
- Zhang J, Sun Z, Han Y, et al. Rnf2 knockdown reduces cell viability and promotes cell cycle arrest in gastric cancer cells. Oncol Lett. 2017;13(5):3817-3822. doi:10.3892/ol.2017.5868

FOXA3: Hepatocyte nuclear factor 3-gamma or Forkhead Box A3 (FOXA3) is a transcription factor that can act as a 'pioneer' factor opening the compacted chromatin for other proteins through interactions with nucleosomal core histones. This in turn, replace linker histones at target enhancer and/or promoter sites. FOXA3 is up-regulated and has oncogenic role in Cholangiocarcinoma. In addition, a similar moiety FOXA2 expression was related with cell proliferation, migration and epithelial mesenchymal transition (EMT) processes of colon cancer. FOXA2 was also downregulated with Aquamin (0.87 fold-change) and no change with calcium (0.99 fold) in our study. Diseases associated with FOXA3 include Maturity-Onset Diabetes Of The Young.

- Chen B, Yu J, Lu L, et al. Upregulated forkhead-box A3 elevates the expression of forkhead-box A1 and forkhead-box A2 to promote metastasis in esophageal cancer. Oncol Lett. 2019;17(5):4351-4360. doi:10.3892/ol.2019.10078
- Thanan R, Kaewlert W, Sakonsinsiri C, et al. Opposing Roles of FoxA1 and FoxA3 in Intrahepatic Cholangiocarcinoma Progression. Int J Mol Sci. 2020;21(5):1796. doi:10.3390/ijms21051796

SNX17: Sorting nexin-17 (SNX17) is a critical regulator of endosomal recycling of numerous surface proteins, including integrins, signaling receptor and channels. SNX17 affects Low-density lipoprotein receptor-related protein 5 (LRP5) and subsequently activate β-catenin protein. SNX17 overexpression increased LRP5 and active β-catenin protein levels in 293T and SW480 cells.

- Osborne DG, Piotrowski JT, Dick CJ, Zhang JS, Billadeau DD. SNX17 affects T cell activation by regulating TCR and integrin recycling. J Immunol. 2015;194(9):4555-4566. doi:10.4049/jimmunol.1402734
- Liu F, Zuo X, Liu Y, et al. Suppression of Membranous LRP5 Recycling, Wnt/β-Catenin Signaling, and Colon Carcinogenesis by 15-LOX-1 Peroxidation of Linoleic Acid in PI3P. Cell Rep. 2020;32(7):108049. doi:10.1016/j.celrep.2020.108049

VAMP3: Vesicle-associated membrane protein 3 (VAMP3) is involved in cargo recognition for clathrin-mediated endocytosis; ER-Phagosome pathway; Retrograde transport at the Trans-Golgi-Network. VAMP3 silencing diminished cell adhesion to laminin but not to fibronectin or collagen. VAMP3-dependent integrin trafficking is crucial in cell migration and cell adhesion to laminin. VAMP3 participates selectively in trafficking of the integrins that function as laminin receptors. Integrin ligation and protein kinases C (PKC) activation are required for migration of colon carcinoma cells. VAMP3 is required for secretion of MMP2 and MMP9 and for efficient invasion by HT-1080 cells in vitro.

- Luftman K, Hasan N, Day P, Hardee D, Hu C. Silencing of VAMP3 inhibits cell migration and integrin-mediated adhesion. Biochem Biophys Res Commun. 2009;380(1):65-70. doi:10.1016/j.bbrc.2009.01.036
- Kean MJ, Williams KC, Skalski M, et al. VAMP3, syntaxin-13 and SNAP23 are involved in secretion of matrix metalloproteinases, degradation of the extracellular matrix and cell invasion. J Cell Sci. 2009;122(Pt 22):4089-4098. doi:10.1242/jcs.052761

CCL15: C-C motif chemokine 15 (CCL15) is a chemotactic factor that attracts T-cells and monocytes, but not neutrophils, eosinophils, or B-cells. Loss of SMAD4 promotes CCL15 expression from CRC cells to recruit CCR1+ myeloid cells, which facilitates primary tumor invasion and liver metastasis of CRC. In our current study, SMAD4 was upregulated with Aquamin (1.25 fold-change), and with calcium (1.11 fold-change).

- Itatani Y, Kawada K, Inamoto S, et al. The Role of Chemokines in Promoting Colorectal Cancer Invasion/Metastasis. Int J Mol Sci. 2016;17(5):643. doi:10.3390/ijms17050643
- Itatani Y, Kawada K, Fujishita T, et al. Loss of SMAD4 from colorectal cancer cells promotes CCL15 expression to recruit CCR1+ myeloid cells and facilitate liver metastasis. Gastroenterology. 2013;145(5):1064-1075.e11. doi:10.1053/j.gastro.2013.07.033
- Inamoto S, Itatani Y, Yamamoto T, et al. Loss of SMAD4 Promotes Colorectal Cancer Progression by Accumulation of Myeloid-Derived Suppressor Cells through the CCL15-CCR1 Chemokine Axis. Clin Cancer Res. 2016;22(2):492-501. doi:10.1158/1078-0432.CCR-15-0726

RPL36: 60S ribosomal protein L36 (RPL36) promotes cell proliferation and G1/S cell cycle progression through a mechanism involving STAT1 that is also downregulated with calcium and Aquamin in the colon biopsies (our current study). Ribosomal proteins (RP) play key roles in the regulation of apoptosis, multidrug resistance and carcinogenesis. RPL36 may be involved in the early stage of hepatocarcinogenesis, and it can be used as an independent and potential prognostic marker for hepatocellular carcinoma.

- Hu YW, Kang CM, Zhao JJ, et al. LncRNA PLAC2 down-regulates RPL36 expression and blocks cell cycle progression in glioma through a mechanism involving STAT1. J Cell Mol Med. 2018;22(1):497-510. doi:10.1111/jcmm.13338
- Song MJ, Jung CK, Park CH, et al. RPL36 as a prognostic marker in hepatocellular carcinoma. Pathol Int. 2011;61(11):638-644. doi:10.1111/j.1440-1827.2011.02716.x

FAM84B: Protein LRATD2 (FAM84B4 or LRATD2) upregulates in colorectal cancer and serves as an oncogenic gene. It is also associated with prostate cancer tumorigenesis and castration-resistant prostate cancer progression. FAM84B4 also induces cell proliferation and metastasis.

- Peng W, Zhang C, Peng J, et al. Lnc-FAM84B-4 acts as an oncogenic lncRNA by interacting with protein hnRNPK to restrain MAPK phosphatases-DUSP1 expression. Cancer Lett. 2020;494:94-106. doi:10.1016/j.canlet.2020.08.036
- Wong N, Gu Y, Kapoor A, et al. Upregulation of FAM84B during prostate cancer progression. Oncotarget. 2017;8(12):19218-19235. doi:10.18632/oncotarget.15168

CREG1: Protein CREG1 may contribute to the transcriptional control of cell growth and differentiation. It is involved in senescence and autophagy in cancer. CREG1 overexpression is closely correlated with KRAS mutation status in NSCLC cells and could be down-regulated by inhibition of KRAS expression. CREG1 is a downstream effector of KRAS in a sub-type of NSCLC cells and a novel candidate biomarker or therapeutic target for KRAS mutant NSCLC. CREG1 is expressed in colorectal cancer cell lines (evaluated by increased protein levels of CREG1 by western blotting).

- Di Bacco A, Gill G. The secreted glycoprotein CREG inhibits cell growth dependent on the mannose-6-phosphate/insulin-like growth factor II receptor. Oncogene. 2003;22(35):5436-5445. doi:10.1038/sj.onc.1206670
- Veal E, Groisman R, Eisenstein M, Gill G. The secreted glycoprotein CREG enhances differentiation of NTERA-2 human embryonal carcinoma cells. Oncogene. 2000;19(17):2120-2128. doi:10.1038/sj.onc.1203529
- Clark DJ, Mei Y, Sun S, Zhang H, Yang AJ, Mao L. Glycoproteomic Approach Identifies KRAS as a Positive Regulator of CREG1 in Non-small Cell Lung Cancer Cells. Theranostics. 2016;6(1):65-77. doi:10.7150/thno.12350

SPATA2: Spermatogenesis-associated protein 2 (SPATA2) is identified as a TNF receptor modulator that is required for TNF-induced inflammation and apoptosis. SPATA2 also has a role in gynecological tumorigenesis.

- Wieser V, Tsibulak I, Degasper C, et al. Tumor necrosis factor receptor modulator spermatogenesis-associated protein 2 is a novel predictor of outcome in ovarian cancer. Cancer Sci. 2019;110(3):1117-1126. doi:10.1111/cas.13955
- Wieser V, Abdel Azim S, Sprung S, et al. TNFα signalling predicts poor prognosis of patients with endometrial cancer. Carcinogenesis. 2020;41(8):1065-1073. doi:10.1093/carcin/bgaa034

UTP18: U3 small nucleolar RNA-associated protein 18 (UTP18) is involved in the major pathway of rRNA processing in the nucleolus and cytosol. UTP18 is identified as one of the genes that could serve as anticancer targets and prognostic markers in colon adenocarcinoma and its subtypes.

- Hu M, Fu X, Si Z, et al. Identification of Differently Expressed Genes Associated With Prognosis and Growth in Colon Adenocarcinoma Based on Integrated Bioinformatics Analysis. Front Genet. 2019;10:1245. doi:10.3389/fgene.2019.01245
- Sun LC, Qian HX. Screening for implicated genes in colorectal cancer using whole-genome gene expression profiling. Mol Med Rep. 2018;17(6):8260-8268. doi:10.3892/mmr.2018.8862

ZNF33B: Zinc finger protein 33B is involved in transcriptional regulation. ZNF331 was reported to be a transcriptional repressor. Methylation of the promoter region of ZNF331 has been found frequently in human esophageal and gastric cancers. The methylation status of ZNF331 can be applied as a prognostic marker of human colorectal cancer (CRC).

- Wang Y, He T, Herman JG, et al. Methylation of ZNF331 is an independent prognostic marker of colorectal cancer and promotes colorectal cancer growth [published correction appears in Clin Epigenetics. 2018 Mar 14;10 :36]. Clin Epigenetics. 2017;9:115. doi:10.1186/s13148-017-0417-4
- Ferbus D, Bovin C, Validire P, Goubin G. The zinc finger protein OZF (ZNF146) is overexpressed in colorectal cancer. J Pathol. 2003;200(2):177-182. doi:10.1002/path.1337
- Binetti M, Lauro A, Vaccari S, Cervellera M, Tonini V. Proteogenomic biomarkers in colorectal cancers: clinical applications. Expert Rev Proteomics. 2020;17(5):355-363. doi:10.1080/14789450.2020.1782202

SUPT4H1: Transcription elongation factor SPT4 (SUPT4H1) is a component of the DRB sensitivity-inducing factor complex (DSIF complex), which regulates mRNA processing and transcription elongation by RNA polymerase II and involved in chromatin remodeling. A knockdown of the expression of RNF43-SUPT4H1 chimeric transcript was found to have a growth-inhibitory effect in colorectal cancer cells. It is also involved in HIV elongation arrest and recovery and formation of the HIV-1 Early Elongation Complex.

- Lee JR, Kwon CH, Choi Y, et al. Transcriptome analysis of paired primary colorectal carcinoma and liver metastases reveals fusion transcripts and similar gene expression profiles in primary carcinoma and liver metastases. BMC Cancer. 2016;16:539. doi:10.1186/s12885-016-2596-3
- Palangat M, Renner DB, Price DH, Landick R. A negative elongation factor for human RNA polymerase II inhibits the anti-arrest transcript-cleavage factor TFIIS. Proc Natl Acad Sci U S A. 2005;102(42):15036-15041. doi:10.1073/pnas.0409405102

CMTM6: CKLF-like MARVEL transmembrane domain-containing protein 6 (CMTM6) is involved in neutrophil degranulation. It is also a master regulator of recycling and plasma membrane expression of PD-L1/CD274, an immune inhibitory ligand critical for immune tolerance to self and antitumor immunity.

- Dorand RD, Petrosiute A, Huang AY. Multifactorial regulators of tumor programmed death-ligand 1 (PD-L1) response. Transl Cancer Res. 2017;6(Suppl 9):S1451-S1454. doi:10.21037/tcr.2017.11.08
- Patsoukis N, Wang Q, Strauss L, Boussiotis VA. Revisiting the PD-1 pathway. Sci Adv. 2020;6(38):eabd2712. doi:10.1126/sciadv.abd2712
